# State-Level Excess Drug Overdose Mortality by Race/Ethnicity in the U.S., 2020–2023: A Population-Scaling Analysis During the COVID-19 Pandemic

**DOI:** 10.64898/2026.01.05.25341762

**Authors:** Faharudeen Alhassan, Hamed Karami, Evangeline Cheng, Sunmi Lee, Isaac C. H. Fung, Robert M. Bohler, Gerardo Chowell

## Abstract

**Purpose:** The COVID-19 pandemic coincided with worsening U.S. drug overdose mortality and widening racial and ethnic disparities. We estimated race/ethnicity–specific excess overdose deaths at the state level during 2020–2023 and examined how these burdens scale with population size.

**Methods:** We analyzed annual overdose deaths from 2014–2023 for five racial/ethnic groups in 47 states and the District of Columbia. Pre-pandemic trends (2014–2019) were fit using a generalized growth model, and deaths exceeding projections in 2020–2023 were classified as excess deaths. Scaling was evaluated by regressing excess and total deaths on 2020 state population in log–log models and relating excess-death rates to social and behavioral indicators.

**Results:** White populations experienced the largest absolute excess, peaking in 2021. Black populations showed minimal excess in 2020 but rose sharply in 2021–2022, nearly matching White deaths by 2023; Hispanic excesses were intermediate. Scaling analyses showed sublinear growth of Black deaths with population, while White and Hispanic deaths scaled approximately proportionally. Within-state rank tests indicated higher excess-death rates for Black than for White or Hispanic populations.

**Conclusions:** Excess-mortality and scaling analyses reveal heterogeneous and inequitable overdose burdens across racial and ethnic groups during the pandemic and can inform equity-focused surveillance and prevention efforts.

## 1. Introduction

The COVID-19 pandemic coincided with a worsening of the U.S. overdose crisis and widening racial and ethnic disparities. Despite a national decline from 2023 to 2024, deaths remain well above pre-pandemic baselines Centers for Disease Control and Prevention (2025). Inequities that were emerging before 2020 intensified; from 2019 to 2020, Black individuals experienced the largest year-over-year increase in overdose mortality (≈44%) Larochelle et al. (2021); Hedegaard et al. (2021). The crisis is geographically uneven, with steep increases and excess mortality concentrated in specific states and regions Chandra et al. (2024); Kiang et al. (2022). At the same time, the substance landscape shifted toward polysubstance involvement, now a hallmark of the epidemic’s “fourth wave” Shover et al. (2020); Friedman and Shover (2023); Friedman et al. (2021). Together, these patterns underscore the need for timely, granular (state-, county-, and group-specific) data to track the evolving overdose crisis and guide policy and practice.

Traditional vital statistics systems often fall short of capturing the full extent of drug-related mortality. A substantial proportion of overdose deaths are misclassified or attributed to ill-defined causes, particularly in cases involving synthetic opioids or mixed drug use, leading to underreporting over time Ruhm (2018). Variations in medical examiner resources, toxicology testing, and death-certificate documentation contribute to reporting inconsistencies across jurisdictions, and these limitations were exacerbated during the COVID-19 pandemic, when strained health systems and shifting cause-of-death attribution (e.g., prioritizing COVID-19 as the underlying cause) further complicated overdose surveillance Centers for Disease Control and Prevention (2020).

Excess-mortality models estimate the number of deaths above an expected baseline, capturing both direct and indirect effects of public health emergencies Mathieu et al. (2020); Leon et al. (2020); Kontis et al. (2020). Widely applied during the COVID-19 pandemic, these models quantified broader mortality impacts beyond confirmed COVID-19 deaths, including the effects of healthcare strain, social disruption, and reduced access to treatment Woolf et al. (2020); Weinberger et al. (2020). Excess mortality does not equate directly to COVID-19 deaths but instead reflects the cumulative toll of the pandemic on vulnerable populations Faust et al. (2021); Haley and Saitz (2020); Slavova et al. (2020); Woolf et al. (2021). Applied to drug-related mortality, excess-mortality frameworks can reveal hidden trends in overdose fatalities where classification is inconsistent or delayed and help identify subgroups and periods with disproportionately elevated risks, informing the targeting of prevention resources and evaluation of sudden social and economic changes Chen and Krieger (2021); Borquez and Martin (2022).

Excess-mortality methods have recently been used to quantify overdose burden during the COVID-19 era by comparing observed deaths to expected baselines. One nationwide analysis estimated that between March 2020 and August 2021 there were approximately 25,000 more fatal drug overdoses than would have been expected in the absence of the pandemic, with the sharpest increases in the South and West Chandra et al. (2024). State-level work has shown similar patterns with even sharper inequities; in California, fatal drug overdoses increased by 44% in 2020 compared to 2019, corresponding to an estimated 2,084 excess overdose deaths concentrated in southern regions and disproportionately borne by structurally marginalized racial and ethnic groups Kiang et al. (2022). A national time-series study covering 2012–2022 further characterized excess overdose deaths at the U.S. population level and documented pandemic-era amplification of overdose mortality, including disparities across subpopulations Zhao et al. (2024). However, no prior work has systematically estimated state-level excess overdose mortality stratified by race/ethnicity through 2023 or examined how overdose burden scales with population size across racial/ethnic groups Kiang et al. (2022); Zhao et al. (2024); Chandra et al. (2024).

This study estimates race/ethnicity–specific excess drug overdose mortality during 2020–2023 by comparing observed overdose deaths to expected counts from a pre-pandemic baseline model. We extend prior work by providing state-level resolution, stratifying results by race/ethnicity, and updating through 2023. Previous analyses have either focused on a single state with detailed demographic breakdowns Kiang et al. (2022) or examined national excess overdose mortality without jointly providing state-by-race estimates Zhao et al. (2024). In addition, we quantify how excess and total overdose deaths scale with state population size across racial/ethnic groups, a novel application of population-scaling analysis to overdose mortality. This allows us to identify whether overdose burden grows proportionally with population or whether particular groups (e.g., Black or Hispanic populations) experience disproportionately high burden in smaller or larger populations.

## 2. Data and Methodology

### 2.1. Data Source

We analyzed annual, state-level counts of drug overdose deaths in the United States from 2014 through 2023, stratified by race/ethnicity. Mortality data were obtained from the CDC WONDER Multiple Cause of Death (MCOD) system Multiple Cause of Death Data on CDC WONDER. To ensure consistent coverage and maximize completeness, we combined two overlapping data series: the Current Final MCOD Data for 1999–2020 and the Current Final MCOD Data for 2018–2023, retaining counts from the 1999–2020 series for overlapping years (2018–2020) to preserve internal consistency in coding conventions, data processing, and population estimates.

Drug overdose deaths were identified using ICD-10 underlying cause-of-death codes X40–X44 (unintentional), X60–X64 (suicide), X85 (homicide), and Y10–Y14 (undetermined), consistent with CDC definitions and prior studies Billock et al. (2023). These categories capture all injury-related drug poisonings regardless of intent. Records associated with other ICD-10 codes were excluded.

Population denominators were derived from CDC Bridged-Race Population Estimates, which provide annual population counts by year, state, race, ethnicity, sex, and age Bridged-Race Population Estimates. We used the 2020 bridged-race population estimates as a fixed reference denominator across all study years to compute overdose mortality and excess-death rates. This provided a consistent reference point across states and racial/ethnic groups. Supplementary Figure S1 illustrates the racial/ethnic composition of each state, highlighting the relative population sizes that underlie the per capita overdose estimates.

To explore ecological correlates of excess overdose mortality, we compiled six state-level socioeconomic and health-behavior indicators (alcohol use, smoking, binge drinking, uninsured status, labor force participation, and poverty) from Global Burden of Disease (GBD), CDC, KFF, BLS/FRED, and the Census SAIPE program. All indicators represent the total state population (not race-specific strata). Detailed variable definitions and measurement details are provided in Supplementary Methods S1–S2 Institute for Health Metrics and Evaluation (IHME) (2025); Centers for Disease Control and Prevention (2024); KFF State Health Facts (2025); Federal Reserve Bank of St. Louis (2025); U.S. Census Bureau (2025).

### 2.2. Data Preparation

We considered five racial/ethnic groups: non-Hispanic White, non-Hispanic Black, Hispanic, non-Hispanic Asian, and non-Hispanic American Indian/Alaska Native (hereafter White, Black, Hispanic, Asian, and Native). Each observation in the analytic dataset includes the state, year (2014–2023), racial/ethnic group, and the corresponding number of reported drug overdose deaths.

Reporting completeness varied across states and racial/ethnic groups. To support reliable intergroup comparisons, we restricted analyses to state–race combinations with complete reporting over 2014–2023. For example, California reported complete data for all five racial/ethnic groups across all years and was fully included, whereas New York lacked complete data for Native American populations in some years, so only four fully reported racial/ethnic groups were analyzed for that state. States such as South Dakota, Rhode Island, and Wyoming, which lacked complete data for one or more groups across the study period, were excluded entirely. Although not a state, the District of Columbia was included due to its complete reporting. In total, the final analysis included 47 states and the District of Columbia.

### 2.3. Modeling Framework for Overdose Mortality

To estimate excess drug overdose mortality, we modeled pre-pandemic trends using two parsimonious approaches: a *Generalized Growth Model (GGM)* and a *Simple Linear Regression (SLR)*. Both were fit to annual state- and race/ethnicity-specific mortality data for 2014–2019 and extrapolated to generate expected deaths for 2020–2023. The GGM captures sub-exponential to exponential growth in cumulative deaths and is well-suited to pre-2020 nonlinear increases in overdose mortality Chowell et al. (2019); Viboud et al. (2016), whereas SLR assumes a constant absolute annual change and serves as a simple benchmark. Detailed model equations, likelihood specification, and implementation are provided in Supplementary Methods S3–S5 Chowell et al. (2024); Bleichrodt et al. (2024).

We compared 2020–2023 projections across state–race strata using Spearman’s rank correlation and assessed calibration over 2014–2019 using mean absolute error (MAE) and mean squared error (MSE). The GGM and SLR projections were highly correlated, and GGM achieved a lower average MAE and MSE, indicating improved fit to the pre-pandemic trajectory (Supplementary Figure S12). We therefore selected the GGM as the primary baseline for estimating excess overdose deaths.

### 2.4. Excess Death Estimation

For each state–race stratum and year, expected deaths in 2020–2023 were obtained from the fitted GGM, and excess deaths were defined as the amount by which observed deaths exceeded expected values. Specifically, we set annual excess deaths to zero when observed deaths were below or equal to expected deaths to focus on positive deviations above the modeled baseline Weinberger et al. (2020). We then summed annual excess deaths over 2020–2023 to obtain cumulative excess counts and calculated corresponding excess-death rates per 100,000 using the fixed 2020 population denominator.

We derived 95% bounds for annual excess deaths using prediction intervals from the GGM projections and aggregated these bounds over 2020–2023 to obtain conservative interval estimates for cumulative excess deaths and rates. This baseline-versus-observed approach has been widely used to assess the impact of major shocks on mortality, including the COVID-19 pandemic Karami et al. (2025); Zhao et al. (2024); Kontis et al. (2020). Further details, including explicit formulas for annual and cumulative excess deaths, interval bounds, and rate calculations, are provided in Supplementary Methods S6.

### 2.5. Summary of Analytical Approach

Our analysis incorporated several complementary procedures to characterize state- and race/ethnicity-specific excess overdose mortality during 2020–2023. First, we computed state-level totals and per capita excess mortality rates with associated uncertainty bounds derived from the expected-death models, presenting these summaries in tables and bar charts. To examine distributional patterns across racial and ethnic groups, we compared excess-mortality rates using log_10_-transformed boxplots for the common three-group overlap (White, Black, Hispanic) and for all available pairwise group intersections. Geographic variation in excess mortality was described using choropleth maps created from square-root–transformed excess-death counts. Population-scaling relationships were evaluated by fitting log–log regressions of excess and total overdose deaths versus 2020 state population size; slope estimates and 95% confidence intervals were obtained separately for each racial/ethnic group and for the aggregate population. Finally, we applied Wilcoxon signed-rank tests to formally compare excess-mortality distributions across racial/ethnic groups for both the three-group overlap and all pairwise intersections, aligning these statistical results with the distributional patterns visualized in the figures.

## 3. Results

Across 2020–2023, states experienced large and heterogeneous increases in excess drug overdose mortality, with consistent racial/ethnic disparities and clear geographic clustering. Patterns in per capita burden, distributional comparisons, and population scaling further revealed systematic differences in how the overdose crisis evolved across groups. Fig. 1, we display representative GGM-based forecasts for White, Black, and Hispanic populations to illustrate the model’s behavior and uncertainty within each group. Below, we summarize these results by racial/ethnic group and analytic domain. Additional detailed results are provided in Supplementary Results S1–S7.

**Figure 1:**
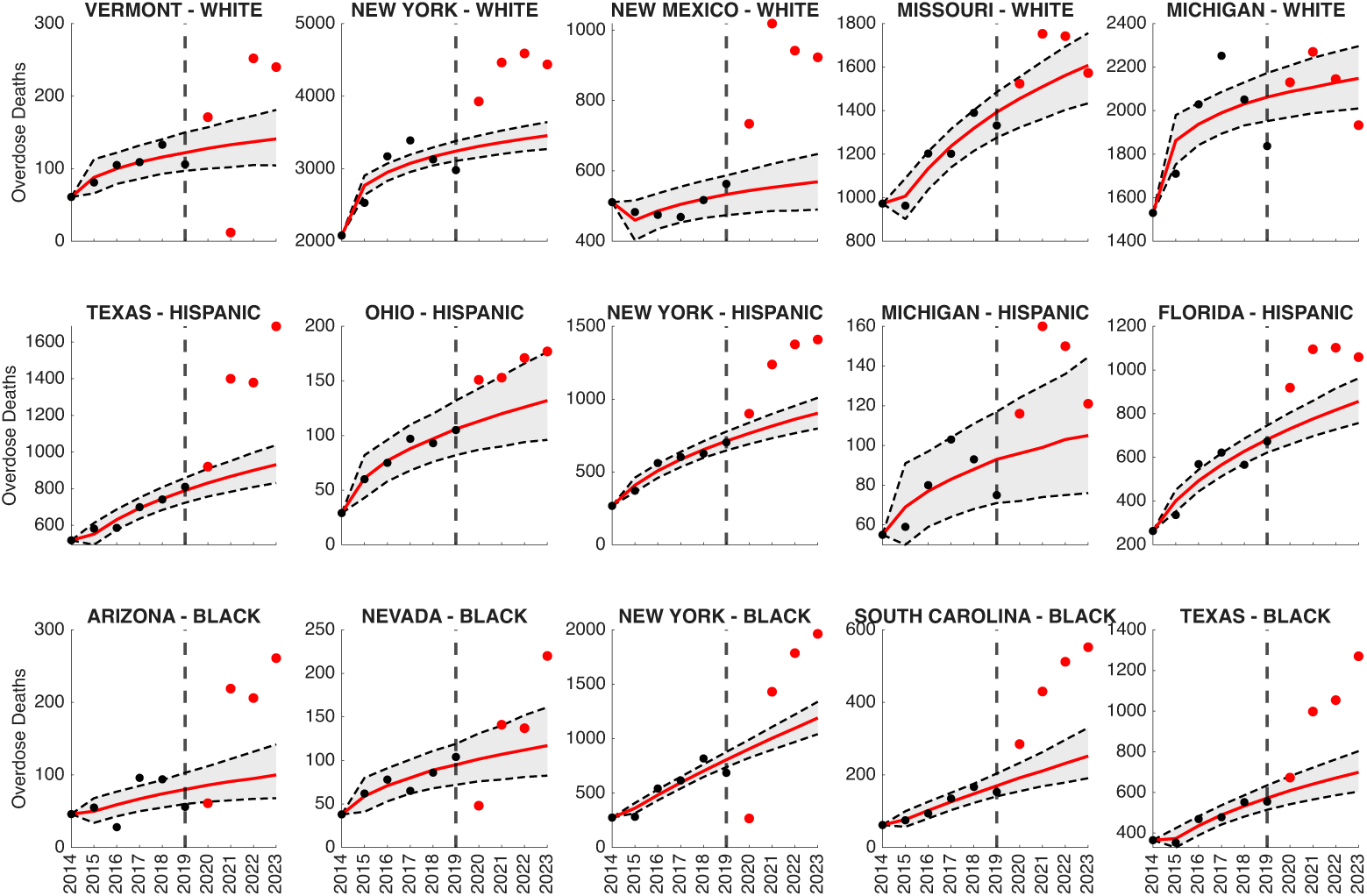
Representative forecasts for the White (first row), Hispanic (second row), and Black (third row) populations using the GGM. The model is calibrated to the pre-pandemic period (2014–2019) and used to generate forecasts for the pandemic period (2020–2023). The vertical black dashed line marks the end of the calibration period. Red curves show the median GGM predictions, and black dashed curves indicate the 95% prediction intervals (PIs). Solid black circles denote observed overdose deaths during calibration, while solid red circles denote observed overdose deaths in the forecast period (2020–2023).

### 3.1. White Populations

White populations experienced the highest absolute number of excess drug overdose deaths from 2020–2023. Across states with complete data, cumulative excess deaths in White populations generally exceeded those in other racial/ethnic groups (Supplementary Figures S3–S5). In the combined three-group comparison (White, Black, Hispanic), White populations carried the largest absolute excess burden, followed by Black and then Hispanic populations. State-level estimates show substantial excess mortality among White residents in large states and in selected Appalachian and Midwestern states, with peaks typically occurring around 2021 and partial declines thereafter (Table 1).

**Table 1.** Annual Excess Drug Overdose Deaths Among White Populations, by State, 2020–2023.

| State | 2020 Estimate [LB, UB] | 2021 Estimate [LB, UB] | 2022 Estimate [LB, UB] | 2023 Estimate [LB, UB] | Total [LB, UB] |
| --- | --- | --- | --- | --- | --- |
| Alabama | 107.0 [42.0, 165.0] | 417.0 [348.0, 476.5] | 467.0 [397.0, 524.0] | 464.0 [388.0, 524.0] | **1455.0 [1175.0, 1689.5]** |
| Alaska | 0.0 [0.0, 0.0] | 0.0 [0.0, 0.0] | 0.0 [0.0, 0.0] | 0.0 [0.0, 0.0] | **0.0 [0.0, 0.0]** |
| Arizona | 495.0 [379.0, 629.0] | 553.0 [419.0, 699.0] | 391.0 [239.5, 555.0] | 369.0 [200.0, 553.0] | **1808.0 [1237.5, 2436.0]** |
| Arkansas | 85.0 [32.0, 132.0] | 141.0 [84.0, 190.0] | 116.0 [56.5, 166.0] | 10.0 [0.0, 64.0] | **352.0 [172.5, 552.0]** |
| California | 2326.0 [2130.0, 2528.0] | 3755.0 [3544.5, 3981.0] | 3575.0 [3340.0, 3821.0] | 3742.5 [3495.0, 4012.0] | **13398.5 [12509.5, 14342.0]** |
| Colorado | 322.0 [229.0, 425.0] | 694.0 [597.0, 812.0] | 579.0 [473.0, 706.0] | 580.0 [463.0, 717.0] | **2175.0 [1762.0, 2660.0]** |
| Connecticut | 59.0 [0.0, 154.0] | 118.0 [10.5, 221.0] | 0.0 [0.0, 100.0] | 0.0 [0.0, 0.0] | **177.0 [10.5, 475.0]** |
| Delaware | 0.0 [0.0, 0.0] | 0.0 [0.0, 0.0] | 0.0 [0.0, 0.0] | 0.0 [0.0, 0.0] | **0.0 [0.0, 0.0]** |
| Florida | 1303.0 [1114.0, 1535.0] | 1635.0 [1425.0, 1889.0] | 1041.0 [806.0, 1327.5] | 398.0 [146.5, 709.0] | **4377.0 [3491.5, 5460.5]** |
| Georgia | 0.0 [0.0, 0.0] | 435.0 [397.0, 468.0] | 556.0 [515.0, 589.0] | 632.0 [589.0, 665.0] | **1623.0 [1501.0, 1722.0]** |
| Hawaii | 5.0 [0.0, 24.0] | 0.0 [0.0, 16.0] | 23.0 [0.0, 43.0] | 29.0 [0.0, 50.0] | **57.0 [0.0, 133.0]** |
| Idaho | 33.0 [0.0, 72.0] | 96.0 [50.0, 136.0] | 92.0 [42.0, 136.0] | 116.0 [61.0, 162.0] | **337.0 [153.0, 506.0]** |
| Illinois | 175.0 [43.0, 330.0] | 147.0 [0.0, 313.5] | 43.0 [0.0, 224.5] | 0.0 [0.0, 35.5] | **365.0 [43.0, 903.5]** |
| Indiana | 391.0 [276.0, 512.0] | 662.0 [536.0, 797.5] | 509.0 [368.5, 654.0] | 93.0 [0.0, 247.5] | **1655.0 [1180.5, 2211.0]** |
| Iowa | 97.0 [48.0, 145.0] | 109.0 [54.0, 158.0] | 92.0 [33.0, 143.0] | 101.0 [37.0, 152.0] | **399.0 [172.0, 598.0]** |
| Kansas | 89.0 [41.0, 139.0] | 246.0 [192.0, 298.0] | 306.0 [247.0, 363.0] | 190.0 [123.0, 248.0] | **831.0 [603.0, 1048.0]** |
| Kentucky | 614.0 [526.0, 695.0] | 829.0 [738.0, 911.0] | 695.0 [602.0, 780.0] | 519.0 [424.0, 606.0] | **2657.0 [2290.0, 2992.0]** |
| Louisiana | 328.0 [240.5, 430.0] | 623.0 [526.0, 740.5] | 473.5 [368.0, 600.0] | 301.0 [184.0, 442.5] | **1725.5 [1318.5, 2213.0]** |
| Maine | 79.0 [27.0, 129.0] | 186.0 [128.0, 238.5] | 267.0 [207.0, 323.0] | 147.0 [80.0, 203.0] | **679.0 [442.0, 893.5]** |
| Maryland | 0.0 [0.0, 79.0] | 0.0 [0.0, 0.0] | 0.0 [0.0, 0.0] | 0.0 [0.0, 0.0] | **0.0 [0.0, 79.0]** |
| Massachusetts | 0.0 [0.0, 0.0] | 69.0 [0.0, 196.0] | 0.0 [0.0, 123.5] | 0.0 [0.0, 0.0] | **69.0 [0.0, 319.5]** |
| Michigan | 43.0 [0.0, 160.0] | 163.0 [27.5, 282.0] | 16.0 [0.0, 146.0] | 0.0 [0.0, 0.0] | **222.0 [27.5, 588.0]** |
| Minnesota | 162.0 [99.5, 225.0] | 325.0 [256.0, 393.0] | 297.0 [224.0, 367.0] | 180.0 [100.0, 256.5] | **964.0 [679.5, 1241.5]** |
| Mississippi | 147.0 [102.0, 186.0] | 283.0 [238.0, 326.0] | 282.0 [233.0, 324.0] | 207.0 [156.0, 251.0] | **919.0 [729.0, 1087.0]** |
| Missouri | 69.0 [0.0, 201.5] | 243.0 [127.0, 391.0] | 180.0 [48.0, 338.5] | 0.0 [0.0, 139.5] | **492.0 [175.0, 1070.5]** |
| Montana | 32.0 [8.0, 53.0] | 33.0 [7.0, 55.0] | 49.0 [22.0, 72.0] | 44.0 [15.0, 66.0] | **158.0 [52.0, 246.0]** |
| Nebraska | 56.0 [24.0, 84.0] | 54.0 [19.0, 81.0] | 42.0 [3.0, 72.0] | 0.0 [0.0, 24.0] | **152.0 [46.0, 261.0]** |
| Nevada | 97.0 [36.0, 154.0] | 189.0 [124.0, 249.0] | 211.0 [142.0, 271.0] | 407.0 [336.0, 466.0] | **904.0 [638.0, 1140.0]** |
| New Hampshire | 0.0 [0.0, 0.0] | 0.0 [0.0, 42.0] | 11.0 [0.0, 61.0] | 0.0 [0.0, 22.0] | **11.0 [0.0, 125.0]** |
| New Jersey | 0.0 [0.0, 0.0] | 0.0 [0.0, 0.0] | 0.0 [0.0, 0.0] | 0.0 [0.0, 0.0] | **0.0 [0.0, 0.0]** |
| New Mexico | 190.0 [131.0, 254.0] | 466.0 [399.0, 533.0] | 381.0 [308.0, 455.0] | 354.0 [275.0, 433.0] | **1391.0 [1113.0, 1675.0]** |
| New York | 619.0 [472.5, 773.0] | 1101.0 [938.0, 1260.5] | 1177.0 [1003.5, 1346.0] | 982.0 [795.0, 1166.5] | **3879.0 [3209.0, 4546.0]** |
| North Carolina | 329.0 [202.5, 472.0] | 793.0 [649.0, 953.5] | 921.0 [764.0, 1090.0] | 539.0 [368.0, 731.5] | **2582.0 [1983.5, 3247.0]** |
| North Dakota | 19.0 [0.0, 37.0] | 20.0 [0.0, 38.0] | 40.0 [13.0, 59.0] | 21.0 [0.0, 42.0] | **100.0 [13.0, 176.0]** |
| Ohio | 358.0 [186.0, 514.0] | 414.0 [231.0, 584.5] | 0.0 [0.0, 166.5] | 0.0 [0.0, 0.0] | **772.0 [417.0, 1265.0]** |
| Oklahoma | 0.0 [0.0, 30.0] | 103.0 [36.0, 161.0] | 273.0 [203.5, 334.0] | 325.0 [252.0, 387.5] | **701.0 [491.5, 912.5]** |
| Oregon | 209.0 [148.0, 268.0] | 0.0 [0.0, 0.0] | 666.0 [596.0, 734.0] | 1041.0 [963.0, 1114.0] | **1916.0 [1707.0, 2116.0]** |
| Pennsylvania | 0.0 [0.0, 17.5] | 0.0 [0.0, 0.0] | 0.0 [0.0, 0.0] | 0.0 [0.0, 0.0] | **0.0 [0.0, 17.5]** |
| Tennessee | 677.0 [555.0, 827.5] | 0.0 [0.0, 0.0] | 1071.0 [917.5, 1265.5] | 838.0 [673.0, 1040.0] | **2586.0 [2145.5, 3133.0]** |
| Texas | 760.0 [616.0, 901.0] | 0.0 [0.0, 0.0] | 1800.0 [1629.0, 1969.5] | 1750.0 [1566.5, 1927.0] | **4310.0 [3811.5, 4797.5]** |
| Utah | 0.0 [0.0, 27.0] | 0.0 [0.0, 0.0] | 0.0 [0.0, 40.0] | 34.0 [0.0, 90.0] | **34.0 [0.0, 157.0]** |
| Vermont | 43.0 [14.0, 71.0] | 0.0 [0.0, 0.0] | 115.0 [79.0, 147.0] | 99.0 [59.0, 135.5] | **257.0 [152.0, 353.5]** |
| Virginia | 357.0 [256.0, 458.0] | 0.0 [0.0, 0.0] | 322.0 [200.0, 442.0] | 197.0 [62.0, 330.0] | **876.0 [518.0, 1230.0]** |
| Washington | 341.0 [251.0, 460.5] | 0.0 [0.0, 0.0] | 1109.0 [1003.0, 1251.5] | 1628.0 [1516.0, 1773.5] | **3078.0 [2770.0, 3485.5]** |
| West Virginia | 388.0 [300.0, 499.0] | 0.0 [0.0, 0.0] | 329.0 [224.0, 467.5] | 372.0 [261.0, 511.0] | **1089.0 [785.0, 1477.5]** |
| Wisconsin | 233.0 [149.0, 325.0] | 0.0 [0.0, 0.0] | 235.5 [132.0, 344.0] | 238.0 [128.0, 355.0] | **706.5 [409.0, 1024.0]** |

### 3.2. Black Populations

Black populations experienced a pronounced increase in excess overdose mortality during 2020–2023, with particularly large surges in certain states. Many states reported little or no excess overdose deaths among Black residents in 2020 but exhibited substantial excess mortality in subsequent years. Across most states with available data, the total excess deaths among Black residents were lower in absolute number than among Whites, yet in several states the excess burden in 2023 was similar in magnitude despite smaller Black populations (Supplementary Figure S3). State-level excess deaths among Black populations displayed wide variation; on the log scale, their central tendency and spread were only modestly lower than for White populations, with the largest cumulative excesses observed in California, Florida, New York, Illinois, and Georgia (Supplementary Figure S6; Table 2).

**Table 2.** Annual Excess Drug Overdose Deaths Among Black Populations, by State, 2020–2023.

| State | 2020 Estimate [LB, UB] | 2021 Estimate [LB, UB] | 2022 Estimate [LB, UB] | 2023 Estimate [LB, UB] | Total [LB, UB] |
| --- | --- | --- | --- | --- | --- |
| Arizona | 0.0 [0.0, 0.0] | 128.0 [97.0, 154.0] | 111.0 [74.0, 139.0] | 161.0 [119.0, 193.0] | **400.0 [290.0, 486.0]** |
| California | 2.0 [0.0, 51.0] | 1023.0 [970.0, 1072.0] | 1022.0 [964.0, 1071.0] | 1243.0 [1180.5, 1291.0] | **3290.0 [3114.5, 3485.0]** |
| Colorado | 0.0 [0.0, 11.0] | 93.0 [68.0, 111.0] | 94.0 [63.0, 114.0] | 138.0 [102.0, 160.0] | **325.0 [233.0, 396.0]** |
| Connecticut | 9.0 [0.0, 33.0] | 169.0 [133.0, 196.0] | 168.0 [125.0, 198.0] | 163.0 [111.0, 197.0] | **509.0 [369.0, 624.0]** |
| District of Columbia | 0.0 [0.0, 0.0] | 232.0 [196.0, 264.0] | 210.0 [172.0, 243.0] | 193.0 [151.0, 228.0] | **635.0 [519.0, 735.0]** |
| Florida | 0.0 [0.0, 0.0] | 603.0 [551.0, 651.0] | 715.5 [659.0, 765.0] | 681.0 [619.0, 733.0] | **1999.5 [1829.0, 2149.0]** |
| Georgia | 0.0 [0.0, 0.0] | 436.0 [397.0, 469.0] | 557.0 [514.0, 590.0] | 633.0 [589.0, 666.0] | **1626.0 [1500.0, 1725.0]** |
| Illinois | 0.0 [0.0, 0.0] | 839.0 [771.5, 904.0] | 938.0 [864.0, 1010.0] | 799.0 [722.0, 873.0] | **2576.0 [2357.5, 2787.0]** |
| Indiana | 0.0 [0.0, 0.0] | 334.0 [300.0, 362.0] | 312.0 [275.0, 342.0] | 238.0 [196.5, 269.0] | **884.0 [771.5, 973.0]** |
| Louisiana | 0.0 [0.0, 0.0] | 443.0 [370.0, 509.0] | 435.0 [347.5, 511.0] | 391.0 [288.0, 478.0] | **1269.0 [1005.5, 1498.0]** |
| Maryland | 0.0 [0.0, 0.0] | 0.0 [0.0, 78.0] | 0.0 [0.0, 0.0] | 0.0 [0.0, 0.0] | **0.0 [0.0, 78.0]** |
| Michigan | 0.0 [0.0, 0.0] | 80.0 [0.0, 170.0] | 0.0 [0.0, 106.0] | 37.5 [0.0, 170.0] | **117.5 [0.0, 446.0]** |
| Minnesota | 0.0 [0.0, 0.0] | 133.0 [96.0, 162.0] | 163.0 [119.0, 195.0] | 192.0 [142.0, 227.0] | **488.0 [357.0, 584.0]** |
| Mississippi | 0.0 [0.0, 10.0] | 114.0 [86.0, 134.0] | 103.0 [70.0, 125.0] | 113.0 [74.0, 137.0] | **330.0 [230.0, 406.0]** |
| Missouri | 0.0 [0.0, 0.0] | 0.0 [0.0, 72.0] | 0.0 [0.0, 34.0] | 0.0 [0.0, 0.0] | **0.0 [0.0, 106.0]** |
| Nevada | 0.0 [0.0, 0.0] | 34.0 [1.0, 63.0] | 25.0 [0.0, 56.0] | 103.0 [59.0, 137.5] | **162.0 [60.0, 256.5]** |
| New Jersey | 0.0 [0.0, 0.0] | 0.0 [0.0, 0.0] | 0.0 [0.0, 0.0] | 0.0 [0.0, 0.0] | **0.0 [0.0, 0.0]** |
| New York | 0.0 [0.0, 0.0] | 427.0 [323.0, 533.0] | 690.0 [564.0, 816.5] | 773.0 [624.0, 921.5] | **1890.0 [1511.0, 2271.0]** |
| North Carolina | 0.0 [0.0, 0.0] | 436.0 [367.0, 501.0] | 535.0 [453.0, 610.0] | 562.0 [466.0, 648.0] | **1533.0 [1286.0, 1759.0]** |
| Ohio | 0.0 [0.0, 0.0] | 113.0 [7.0, 211.0] | 93.0 [0.0, 211.0] | 0.0 [0.0, 121.0] | **206.0 [7.0, 543.0]** |
| Oklahoma | 0.0 [0.0, 0.0] | 62.0 [39.0, 80.0] | 79.0 [53.0, 98.0] | 103.0 [73.0, 123.0] | **244.0 [165.0, 301.0]** |
| Pennsylvania | 211.0 [135.0, 282.0] | 285.0 [197.0, 366.0] | 362.0 [260.0, 454.0] | 364.5 [243.5, 466.0] | **1222.5 [835.5, 1568.0]** |
| South Carolina | 93.0 [52.0, 130.0] | 219.0 [167.0, 261.0] | 280.0 [215.0, 333.0] | 300.0 [222.0, 361.0] | **892.0 [656.0, 1085.0]** |
| Tennessee | 218.0 [161.0, 267.0] | 390.0 [322.0, 447.0] | 453.0 [362.0, 519.5] | 370.0 [262.0, 451.0] | **1431.0 [1107.0, 1684.5]** |
| Texas | 64.0 [0.0, 131.0] | 357.0 [276.0, 433.0] | 383.0 [291.0, 468.0] | 571.0 [467.0, 666.0] | **1375.0 [1034.0, 1698.0]** |
| Virginia | 207.0 [152.0, 255.0] | 352.0 [289.0, 408.0] | 376.0 [302.0, 439.0] | 429.0 [341.0, 499.0] | **1364.0 [1084.0, 1601.0]** |
| Washington | 37.0 [16.0, 55.0] | 139.0 [116.0, 157.0] | 194.0 [170.0, 213.5] | 328.0 [302.0, 347.0] | **698.0 [604.0, 772.5]** |
| Wisconsin | 44.0 [4.0, 78.0] | 154.0 [107.0, 193.0] | 165.0 [112.0, 209.0] | 175.0 [112.0, 224.0] | **538.0 [335.0, 704.0]** |

### 3.3. Hispanic Populations

Hispanic populations also endured notable excess overdose mortality from 2020–2023, though in most states the absolute numbers were lower than in White or Black populations. In aggregate, total excess deaths among Hispanics were substantially below those of Whites, making Hispanics the lowest of the three groups in cumulative 2020–2023 burden in the three-group comparison (Supplementary Figures S4–S5). Many states showed relatively modest Hispanic excess deaths, and some exhibited little or no measurable surge; however, states with large Hispanic populations, such as California, New York, and Texas, experienced sizable increases (Table 3; Supplementary Figure S6).

**Table 3.** Annual Excess Drug Overdose Deaths Among Hispanic Populations, by State, 2020–2023.

| State | 2020 Estimate [LB, UB] | 2021 Estimate [LB, UB] | 2022 Estimate [LB, UB] | 2023 Estimate [LB, UB] | Total [LB, UB] |
| --- | --- | --- | --- | --- | --- |
| Arizona | 148.0 [64.0, 225.0] | 89.0 [0.0, 196.0] | 0.0 [0.0, 5.5] | 0.0 [0.0, 0.0] | **237.0 [64.0, 426.5]** |
| California | 881.0 [739.5, 1008.0] | 1116.0 [920.5, 1283.0] | 1057.0 [775.0, 1270.0] | 948.0 [575.5, 1232.5] | **4002.0 [3010.5, 4793.5]** |
| Colorado | 117.0 [69.0, 163.0] | 215.0 [156.0, 266.0] | 183.0 [115.5, 242.5] | 187.0 [107.0, 256.0] | **702.0 [447.5, 927.5]** |
| Connecticut | 14.0 [0.0, 52.0] | 32.0 [0.0, 78.0] | 50.0 [0.0, 107.0] | 0.0 [0.0, 47.0] | **96.0 [0.0, 284.0]** |
| Florida | 188.0 [114.0, 260.0] | 319.0 [236.0, 398.0] | 285.0 [186.0, 374.0] | 203.0 [97.0, 302.0] | **995.0 [633.0, 1334.0]** |
| Illinois | 137.0 [90.5, 181.0] | 132.0 [78.0, 183.0] | 137.0 [76.0, 193.0] | 96.0 [27.0, 159.0] | **502.0 [271.5, 716.0]** |
| Indiana | 10.0 [0.0, 31.0] | 41.0 [5.0, 67.0] | 19.0 [0.0, 50.0] | 42.0 [0.0, 78.0] | **112.0 [5.0, 226.0]** |
| Massachusetts | 0.0 [0.0, 43.5] | 7.0 [0.0, 61.0] | 42.0 [0.0, 103.5] | 13.0 [0.0, 81.0] | **62.0 [0.0, 289.0]** |
| Michigan | 20.0 [0.0, 44.0] | 61.0 [30.0, 86.0] | 47.0 [14.0, 75.0] | 16.0 [0.0, 45.0] | **144.0 [44.0, 250.0]** |
| Nevada | 57.0 [31.0, 80.0] | 80.0 [49.0, 105.0] | 78.0 [41.0, 104.0] | 125.0 [82.0, 153.0] | **340.0 [203.0, 442.0]** |
| New Jersey | 0.0 [0.0, 8.0] | 0.0 [0.0, 30.0] | 0.0 [0.0, 21.0] | 0.0 [0.0, 0.0] | **0.0 [0.0, 59.0]** |
| New Mexico | 120.5 [70.0, 165.0] | 303.0 [248.0, 351.0] | 237.0 [173.0, 289.0] | 234.0 [166.0, 290.0] | **894.5 [657.0, 1095.0]** |
| New York | 135.0 [62.0, 208.5] | 423.0 [339.0, 506.5] | 514.0 [422.0, 610.0] | 506.0 [400.0, 611.0] | **1578.0 [1223.0, 1936.0]** |
| North Carolina | 38.0 [12.0, 58.0] | 90.0 [57.0, 112.5] | 125.0 [85.0, 152.0] | 136.0 [89.0, 165.0] | **389.0 [243.0, 487.5]** |
| Ohio | 38.0 [8.0, 64.0] | 33.0 [0.0, 63.0] | 45.0 [5.0, 77.0] | 45.0 [0.5, 81.0] | **161.0 [13.5, 285.0]** |
| Pennsylvania | 0.0 [0.0, 0.0] | 5.0 [0.0, 72.0] | 0.0 [0.0, 0.0] | 0.0 [0.0, 55.0] | **5.0 [0.0, 127.0]** |
| Texas | 89.0 [11.0, 163.0] | 533.0 [449.0, 617.0] | 479.0 [382.0, 570.0] | 755.0 [648.5, 855.0] | **1856.0 [1490.5, 2205.0]** |
| Utah | 6.0 [0.0, 22.0] | 27.0 [2.0, 45.0] | 6.0 [0.0, 27.0] | 21.0 [0.0, 44.0] | **60.0 [2.0, 138.0]** |
| Washington | 41.0 [17.0, 63.0] | 110.0 [84.0, 135.0] | 164.0 [135.0, 190.0] | 263.0 [231.0, 289.0] | **578.0 [467.0, 677.0]** |
| Wisconsin | 26.0 [2.0, 45.5] | 41.0 [12.0, 63.0] | 67.0 [32.0, 91.0] | 76.0 [34.0, 102.0] | **210.0 [80.0, 301.5]** |

### 3.4. Asian Populations

Asian populations experienced the smallest excess overdose mortality impact in the study, and data were available for only a small number of states. In many states with smaller Asian communities, no significant excess deaths were detected during 2020–2023. In the few states with sufficient data, excess counts remained modest, with detectable increases mainly in California and New York (Supplementary Figure S6; Table 4). Overall, Asian Americans had low excess overdose mortality over 2020–2023, further constrained by limited data coverage.

**Table 4.** Annual Excess Drug Overdose Deaths Among Asian Populations, by State, 2020–2023.

| State | 2020 Estimate [LB, UB] | 2021 Estimate [LB, UB] | 2022 Estimate [LB, UB] | 2023 Estimate [LB, UB] | Total [LB, UB] |
| --- | --- | --- | --- | --- | --- |
| California | 107.0 [66.0, 145.0] | 117.0 [73.0, 158.0] | 227.0 [177.0, 270.0] | 211.0 [155.0, 255.0] | **662.0 [471.0, 828.0]** |
| Hawaii | 0.0 [0.0, 0.0] | 0.0 [0.0, 0.0] | 0.0 [0.0, 0.0] | 0.0 [0.0, 0.0] | **0.0 [0.0, 0.0]** |
| New York | 7.0 [0.0, 24.0] | 23.0 [0.0, 41.0] | 37.0 [11.0, 56.0] | 15.0 [0.0, 35.0] | **82.0 [11.0, 156.0]** |

### 3.5. Native American Populations

Excess overdose deaths among Native American populations showed heterogeneous patterns across states, with a few states experiencing pronounced surges while others saw minimal impact. Nationally, the total excess deaths in this group were much lower in absolute count than those in White, Black, or Hispanic populations, reflecting both smaller population size and more limited geographic coverage. Nevertheless, certain states with substantial Native American communities (e.g., in the West and Plains) registered notable and, in some cases, increasing excess burdens over 2020–2023 (Supplementary Figure S6; Table 5).

**Table 5.** Annual Excess Drug Overdose Deaths Among Native Populations, by State, 2020–2023.

| State | 2020 Estimate [LB, UB] | 2021 Estimate [LB, UB] | 2022 Estimate [LB, UB] | 2023 Estimate [LB, UB] | Total [LB, UB] |
| --- | --- | --- | --- | --- | --- |
| Minnesota | 38.0 [13.0, 57.0] | 76.0 [47.0, 98.0] | 76.0 [42.0, 100.5] | 73.0 [32.0, 101.0] | **263.0 [134.0, 356.5]** |
| New Mexico | 26.0 [11.0, 39.0] | 75.0 [58.0, 88.0] | 78.0 [59.0, 92.0] | 87.0 [67.0, 102.0] | **266.0 [195.0, 321.0]** |
| North Carolina | 35.0 [7.0, 61.0] | 16.0 [0.0, 57.0] | 0.0 [0.0, 52.0] | 0.0 [0.0, 45.0] | **51.0 [7.0, 215.0]** |
| Oklahoma | 0.0 [0.0, 13.0] | 49.0 [22.0, 69.0] | 85.0 [56.0, 106.0] | 94.0 [64.0, 115.0] | **228.0 [142.0, 303.0]** |
| Washington | 23.0 [1.0, 41.0] | 81.0 [56.0, 100.0] | 104.0 [76.0, 125.0] | 132.0 [101.0, 154.0] | **340.0 [234.0, 420.0]** |

### 3.6. Scaling of Excess and Total Overdose Deaths with State Population

To assess how both excess and overall overdose mortality scale with state population size, we generated log_10_–log_10_ scatterplots of deaths versus 2020 population for Black, White, Hispanic, and all-race populations, including only states with available data. The first set of panels (Figure 2) shows total overdose deaths plotted against state population, while the second set of panels (Figure 3) shows excess overdose deaths versus population. In each panel, the red line is an ordinary least-squares fit on log-transformed values; panel annotations report the fitted slope *m* with its 95% confidence interval, adjusted *R*^2^, root mean squared error (RMSE), and sample size *N* .

**Figure 2.**
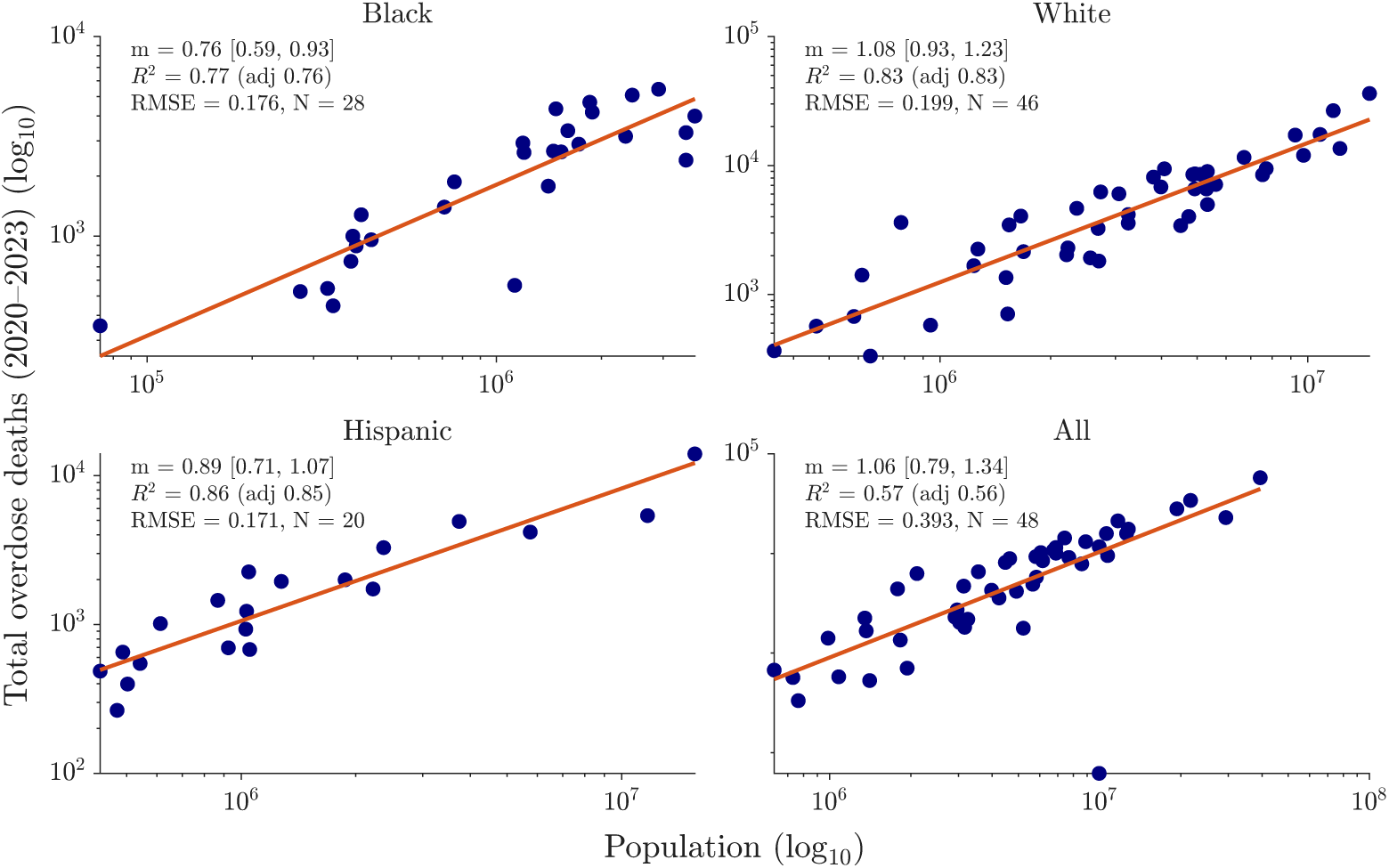
: Relationship between drug overdose deaths and state population size (2020). Each panel shows a log10–log10 scatterplot of overdose deaths versus total population for Black (top left), White (top right), Hispanic (bottom left), and all races combined (bottom right). The solid red line is the OLS fit in log space; panel annotations report the scaling slope *m* with its 95% CI, *R*^2^ (adjusted), RMSE, and *N* . A slope of *m*=1 indicates proportional (isometric) scaling, *m<*1 sublinear, and *m>*1 superlinear.

**Figure 3.**
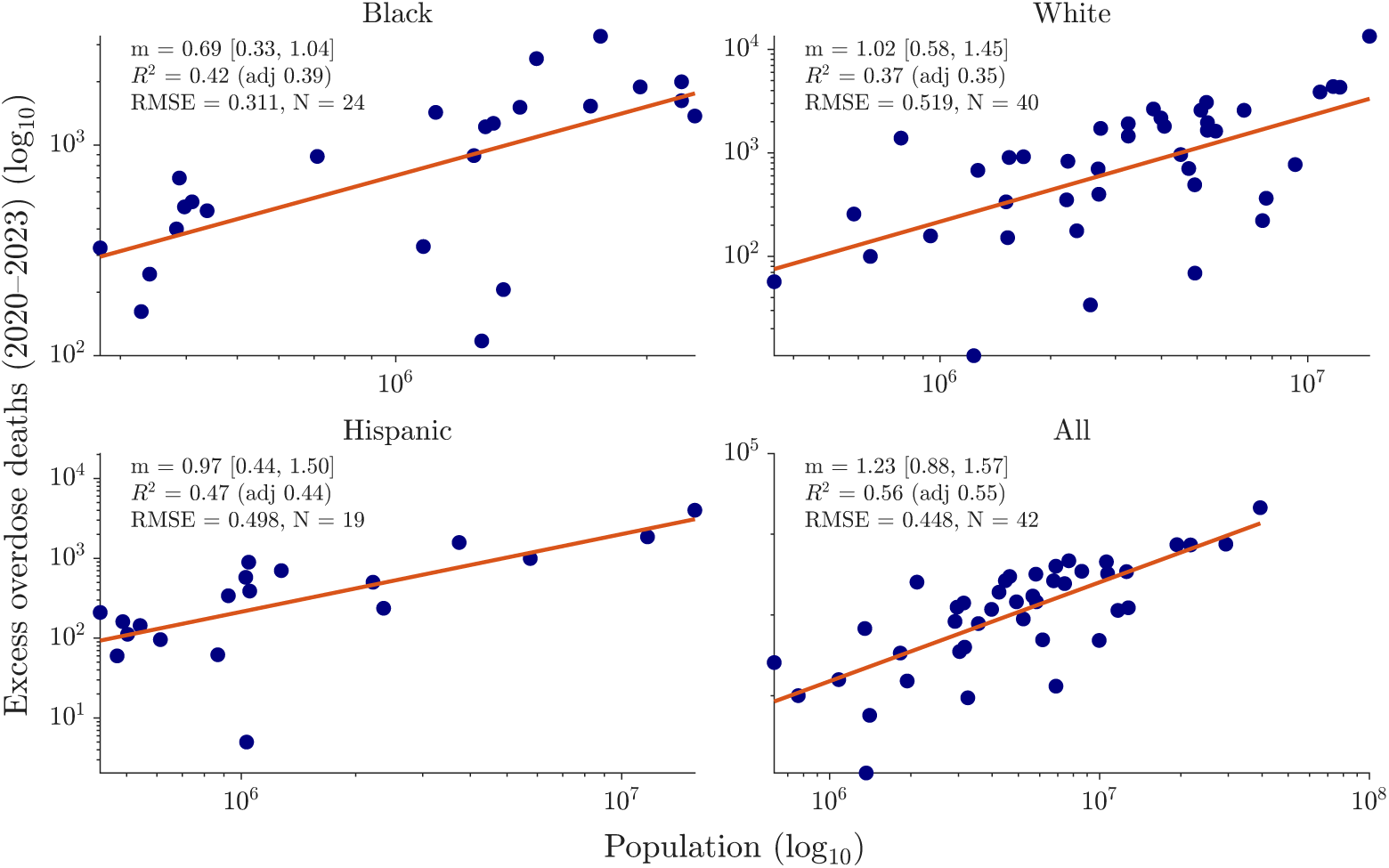
: Relationship between excess drug overdose deaths and state population size (2020). Each panel shows a log10–log10 scatterplot of excess overdose deaths versus total population for Black (top left), White (top right), Hispanic (bottom left), and all races combined (bottom right). The solid red line is the OLS fit in log space; panel annotations report the scaling slope *m* with its 95% CI, *R*^2^ (adjusted), RMSE, and *N* . A slope of *m*=1 indicates proportional (isometric) scaling, *m<*1 sublinear, and *m>*1 superlinear.

#### 3.6.1. Black populations

Figures 2 (total deaths) and 3 (excess deaths) indicate sublinear scaling for total deaths and suggest (but do not conclusively establish) sublinear scaling for excess deaths among Black residents. Excess deaths rise with a slope

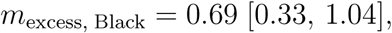

whose confidence interval slightly overlaps 1, whereas total deaths have

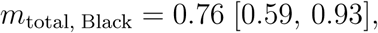

with the entire interval below 1. Thus, total overdose deaths scale sublinearly, and excess deaths are consistent with sublinearity but not definitive.

### 3.6.2. White populations

For White residents, Figures 3 and 2 show point estimates slightly above proportionality. Excess deaths scale at

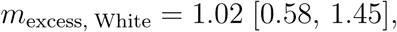

and total deaths at

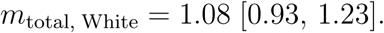

Because both confidence intervals include 1, these patterns are best described as approximately proportional. The point estimates exceed 1—consistent with possible superlinearity—but the evidence is not conclusive.

#### 3.6.3. Hispanic populations

Among Hispanic residents, excess deaths scale nearly one-for-one with population size,

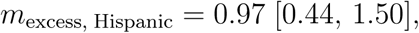

and total deaths show a similar pattern,

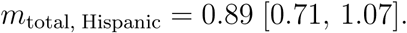

Because both intervals include 1, there is no strong evidence of sub- or superlinearity; a doubling of the Hispanic population roughly doubles both excess and total overdose deaths within statistical uncertainty.

#### 3.6.4. All races combined

When all races are pooled, excess deaths have a point estimate above proportionality,

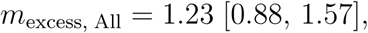

whereas total deaths are nearly proportional,

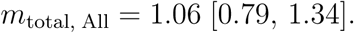

Because the excess-death interval overlaps 1, evidence for superlinear scaling at the aggregate level is suggestive but not definitive.

## 4. Excess Mortality Rate Per 100,000 Population

Figure 5 shows marked geographic heterogeneity in excess overdose mortality rates (2020–2023), with pronounced differences by race/ethnicity. Among White residents, elevated rates cluster in the Southwest (notably New Mexico), extend along the West Coast (California), and appear in parts of Appalachia and the Lower Mississippi Valley. For Black populations, higher rates are concentrated in a Midwest corridor with additional pockets in the West; in contrast, much of the Southeast exhibits comparatively lower rates. The Hispanic panel shows a prominent peak in New Mexico, with moderate levels in parts of the Mountain West and Northeast. The largest rates observed in any panel occur among American Indian/Alaska Native (AI/AN) populations, concentrated in Minnesota and Washington, with additional elevations in New Mexico and Oklahoma. Excess mortality among the Asian population is primarily detectable in California and New York.

Some states appear in gray, indicating missing race-specific data. These gaps—especially for AI/AN and Asian populations—reduce coverage and should be interpreted with caution. Note that each panel uses a different color scale, so visual comparisons should rely on the numerical rates reported in Tables 6–10, rather than color intensity.

**Table 6.** Annual Excess Drug Overdose Death Rates Among White Populations, by State, 2020–2023.

| State | Rate 2020 [LB, UB] | Rate 2021 [LB, UB] | Rate 2022 [LB, UB] | Rate 2023 [LB, UB] | Total [LB, UB] |
| --- | --- | --- | --- | --- | --- |
| Alabama | 3.3 [1.3, 5.1] | 12.8 [10.7, 14.7] | 14.4 [12.2, 16.1] | 14.3 [11.9, 16.1] | **44.8 [36.1, 52]** |
| Alaska | 0 [0, 0] | 0 [0, 0] | 0 [0, 0] | 0 [0, 0] | **0 [0, 0]** |
| Arizona | 12.1 [9.3, 15.4] | 13.6 [10.3, 17.2] | 9.6 [5.9, 13.6] | 9.1 [4.9, 13.6] | **44.4 [30.4, 59.8]** |
| Arkansas | 3.8 [1.4, 6] | 6.4 [3.8, 8.6] | 5.2 [2.6, 7.5] | 0.5 [0, 2.9] | **15.9 [7.8, 25]** |
| California | 15.8 [14.5, 17.2] | 25.5 [24.1, 27] | 24.3 [22.7, 26] | 25.4 [23.7, 27.3] | **91 [85, 97.4]** |
| Colorado | 8.1 [5.7, 10.7] | 17.4 [15, 20.4] | 14.5 [11.9, 17.7] | 14.5 [11.6, 18] | **54.5 [44.2, 66.7]** |
| Connecticut | 2.5 [0, 6.5] | 5 [0.4, 9.4] | 0 [0, 4.2] | 0 [0, 0] | **7.5 [0.4, 20.2]** |
| Delaware | 0 [0, 0] | 0 [0, 0] | 0 [0, 0] | 0 [0, 0] | **0 [0, 0]** |
| Florida | 11.1 [9.5, 13.1] | 13.9 [12.1, 16.1] | 8.9 [6.9, 11.3] | 3.4 [1.2, 6] | **37.3 [29.8, 46.5]** |
| Georgia | 0 [0, 0] | 7.7 [7.1, 8.3] | 9.9 [9.2, 10.5] | 11.2 [10.5, 11.8] | **28.9 [26.7, 30.6]** |
| Hawaii | 1.4 [0, 6.8] | 0 [0, 4.5] | 6.5 [0, 12.1] | 8.2 [0, 14.1] | **16.1 [0, 37.5]** |
| Idaho | 2.2 [0, 4.8] | 6.3 [3.3, 9] | 6.1 [2.8, 9] | 7.7 [4, 10.7] | **22.3 [10.1, 33.5]** |
| Illinois | 2.3 [0.6, 4.3] | 1.9 [0, 4.1] | 0.6 [0, 2.9] | 0 [0, 0.5] | **4.7 [0.6, 11.7]** |
| Indiana | 7.3 [5.2, 9.6] | 12.4 [10, 15] | 9.5 [6.9, 12.3] | 1.7 [0, 4.6] | **31 [22.1, 41.5]** |
| Iowa | 3.6 [1.8, 5.4] | 4 [2, 5.8] | 3.4 [1.2, 5.3] | 3.7 [1.4, 5.6] | **14.8 [6.4, 22.1]** |
| Kansas | 4 [1.8, 6.2] | 11 [8.6, 13.4] | 13.7 [11.1, 16.3] | 8.5 [5.5, 11.1] | **37.3 [27.1, 47]** |
| Kentucky | 16.1 [13.8, 18.3] | 21.8 [19.4, 23.9] | 18.3 [15.8, 20.5] | 13.6 [11.1, 15.9] | **69.8 [60.2, 78.6]** |
| Louisiana | 12 [8.8, 15.7] | 22.8 [19.2, 27.1] | 17.3 [13.5, 21.9] | 11 [6.7, 16.2] | **63.1 [48.2, 80.9]** |
| Maine | 6.2 [2.1, 10.2] | 14.7 [10.1, 18.8] | 21.1 [16.3, 25.5] | 11.6 [6.3, 16] | **53.6 [34.9, 70.5]** |
| Maryland | 0 [0, 2.6] | 0 [0, 0] | 0 [0, 0] | 0 [0, 0] | **0 [0, 2.6]** |
| Massachusetts | 0 [0, 0] | 1.4 [0, 4] | 0 [0, 2.5] | 0 [0, 0] | **1.4 [0, 6.5]** |
| Michigan | 0.6 [0, 2.1] | 2.2 [0.4, 3.7] | 0.2 [0, 1.9] | 0 [0, 0] | **2.9 [0.4, 7.8]** |
| Minnesota | 3.6 [2.2, 5] | 7.2 [5.7, 8.7] | 6.6 [5, 8.1] | 4 [2.2, 5.7] | **21.4 [15.1, 27.5]** |
| Mississippi | 8.7 [6, 11] | 16.8 [14.1, 19.3] | 16.7 [13.8, 19.2] | 12.3 [9.2, 14.9] | **54.5 [43.2, 64.4]** |
| Missouri | 1.4 [0, 4.1] | 4.9 [2.6, 7.9] | 3.7 [1, 6.9] | 0 [0, 2.8] | **10 [3.6, 21.7]** |
| Montana | 3.4 [0.8, 5.6] | 3.5 [0.7, 5.8] | 5.2 [2.3, 7.6] | 4.7 [1.6, 7] | **16.8 [5.5, 26.1]** |
| Nebraska | 3.7 [1.6, 5.5] | 3.5 [1.2, 5.3] | 2.7 [0.2, 4.7] | 0 [0, 1.6] | **10 [3, 17.1]** |
| Nevada | 6.3 [2.3, 10] | 12.3 [8, 16.2] | 13.7 [9.2, 17.6] | 26.4 [21.8, 30.2] | **58.6 [41.4, 74]** |
| New Hampshire | 0 [0, 0] | 0 [0, 3.4] | 0.9 [0, 4.9] | 0 [0, 1.8] | **0.9 [0, 10.1]** |
| New Jersey | 0 [0, 0] | 0 [0, 0] | 0 [0, 0] | 0 [0, 0] | **0 [0, 0]** |
| New Mexico | 24.3 [16.7, 32.4] | 59.5 [50.9, 68] | 48.6 [39.3, 58.1] | 45.2 [35.1, 55.3] | **177.5 [142.1, 213.8]** |
| New York | 5.7 [4.4, 7.2] | 10.2 [8.7, 11.7] | 10.9 [9.3, 12.5] | 9.1 [7.4, 10.8] | **35.9 [29.7, 42.1]** |
| North Carolina | 4.9 [3, 7] | 11.8 [9.7, 14.2] | 13.7 [11.4, 16.2] | 8 [5.5, 10.9] | **38.5 [29.6, 48.4]** |
| North Dakota | 2.9 [0, 5.7] | 3.1 [0, 5.9] | 6.2 [2, 9.1] | 3.2 [0, 6.5] | **15.5 [2, 27.2]** |
| Ohio | 3.9 [2, 5.6] | 4.5 [2.5, 6.3] | 0 [0, 1.8] | 0 [0, 0] | **8.3 [4.5, 13.7]** |
| Oklahoma | 0 [0, 1.1] | 3.8 [1.3, 6] | 10.1 [7.6, 12.4] | 12.1 [9.4, 14.4] | **26 [18.3, 33.9]** |
| Oregon | 6.4 [4.6, 8.2] | 0 [0, 0] | 20.5 [18.3, 22.6] | 32 [29.6, 34.3] | **58.9 [52.5, 65.1]** |
| Pennsylvania | 0 [0, 0.2] | 0 [0, 0] | 0 [0, 0] | 0 [0, 0] | **0 [0, 0.2]** |
| Tennessee | 13.2 [10.8, 16.2] | 0 [0, 0] | 20.9 [17.9, 24.7] | 16.4 [13.1, 20.3] | **50.5 [41.9, 61.2]** |
| Texas | 6.2 [5, 7.4] | 0 [0, 0] | 14.7 [13.3, 16.1] | 14.3 [12.8, 15.8] | **35.2 [31.2, 39.2]** |
| Utah | 0 [0, 1.1] | 0 [0, 0] | 0 [0, 1.6] | 1.3 [0, 3.5] | **1.3 [0, 6.1]** |
| Vermont | 7.4 [2.4, 12.2] | 0 [0, 0] | 19.7 [13.5, 25.2] | 17 [10.1, 23.2] | **44.1 [26.1, 60.6]** |
| Virginia | 6.7 [4.8, 8.6] | 0 [0, 0] | 6.2 [4.2, 8.8] | 3.7 [1.2, 6.2] | **36.8 [24.4, 50.7]** |
| West Virginia | 23.4 [18.1, 30.1] | 0 [0, 0] | 19.8 [13.5, 28.2] | 22.4 [13.5, 28.2] | **65.7 [47.3, 89.1]** |
| Washington | 6.4 [4.7, 8.7] | 0 [0, 0] | 20.9 [18.9, 23.6] | 30.7 [28.6, 33.4] | **58 [52.2, 65.7]** |
| Wisconsin | 4.9 [3.1, 6.8] | 0 [0, 0] | 5 [2.8, 7.2] | 5 [2.7, 7.5] | **14.9 [8.6, 21.6]** |

**Table 7.** Annual Excess Drug Overdose Death Rates Among Black Populations, by State, 2020–2023.

| State | Rate 2020 [LB, UB] | Rate 2021 [LB, UB] | Rate 2022 [LB, UB] | Rate 2023 [LB, UB] | Total [LB, UB] |
| --- | --- | --- | --- | --- | --- |
| Arizona | 0 [0, 0] | 33.4 [25.3, 40.2] | 29 [19.3, 36.3] | 42 [31, 50.4] | **104.4 [75.7, 126.8]** |
| California | 0.1 [0, 2.1] | 41.8 [39.6, 43.8] | 41.7 [39.4, 43.7] | 50.8 [48.2, 52.7] | **134.3 [127.2, 142.3]** |
| Colorado | 0 [0, 4] | 33.9 [24.8, 40.4] | 34.2 [22.9, 41.5] | 50.3 [37.2, 58.3] | **118.4 [84.9, 144.2]** |
| Connecticut | 2.3 [0, 8.3] | 42.6 [33.5, 49.4] | 42.3 [31.5, 49.9] | 41.1 [28, 49.6] | **128.3 [93, 157.2]** |
| District of Columbia | 0 [0, 0] | 72.2 [61, 82.1] | 65.3 [53.5, 75.6] | 60 [47, 70.9] | **197.6 [161.5, 228.7]** |
| Florida | 0 [0, 0] | 17.3 [15.8, 18.6] | 20.5 [18.9, 21.9] | 19.5 [17.7, 21] | **57.3 [52.4, 61.6]** |
| Georgia | 0 [0, 0] | 12.5 [11.4, 13.4] | 16 [14.7, 16.9] | 18.1 [16.9, 19.1] | **46.6 [43, 49.4]** |
| Illinois | 0 [0, 0] | 45.3 [41.7, 48.8] | 50.7 [46.7, 54.6] | 43.2 [39, 47.2] | **139.2 [127.4, 150.6]** |
| Indiana | 0 [0, 0] | 47.1 [42.3, 51] | 44 [38.8, 48.2] | 33.5 [27.7, 37.9] | **124.6 [108.7, 137.1]** |
| Louisiana | 0 [0, 0] | 28.9 [24.1, 33.2] | 28.4 [22.7, 33.3] | 25.5 [18.8, 31.2] | **82.8 [65.6, 97.7]** |
| Maryland | 0 [0, 0] | 0 [0, 4.1] | 0 [0, 0] | 0 [0, 0] | **0 [0, 4.1]** |
| Michigan | 0 [0, 0] | 5.5 [0, 11.7] | 0 [0, 7.3] | 2.6 [0, 11.7] | **8.1 [0, 30.6]** |
| Minnesota | 0 [0, 0] | 30.3 [21.9, 37] | 37.2 [27.2, 44.5] | 43.8 [32.4, 51.8] | **111.3 [81.5, 133.3]** |
| Mississippi | 0 [0, 0.9] | 10.1 [7.6, 11.9] | 9.1 [6.2, 11.1] | 10 [6.6, 12.1] | **29.2 [20.4, 36]** |
| Missouri | 0 [0, 0] | 0 [0, 9.5] | 0 [0, 4.5] | 0 [0, 0] | **0 [0, 14]** |
| Nevada | 0 [0, 0] | 10.3 [0.3, 19.2] | 7.6 [0, 17] | 31.3 [18, 41.8] | **49.3 [18.3, 78.1]** |
| New Jersey | 0 [0, 0] | 0 [0, 0] | 0 [0, 0] | 0 [0, 0] | **0 [0, 0]** |
| New York | 0 [0, 0] | 14.7 [11.1, 18.3] | 23.7 [19.4, 28] | 26.5 [21.4, 31.6] | **64.9 [51.9, 77.9]** |
| North Carolina | 0 [0, 0] | 18.6 [15.7, 21.4] | 22.8 [19.3, 26] | 24 [19.9, 27.6] | **65.4 [54.9, 75]** |
| Ohio | 0 [0, 0] | 7.1 [0.4, 13.2] | 5.8 [0, 13.2] | 0 [0, 7.6] | **12.9 [0.4, 33.9]** |
| Oklahoma | 0 [0, 0] | 18.2 [11.4, 23.5] | 23.2 [15.5, 28.8] | 30.2 [21.4, 36.1] | **71.6 [48.4, 88.3]** |
| Pennsylvania | 14.3 [9.1, 19] | 19.2 [13.3, 24.7] | 24.5 [17.6, 30.7] | 24.6 [16.4, 31.5] | **82.6 [56.4, 105.9]** |
| South Carolina | 6.6 [3.7, 9.2] | 15.6 [11.9, 18.5] | 19.9 [15.3, 23.7] | 21.3 [15.8, 25.6] | **63.4 [46.6, 77.1]** |
| Tennessee | 18.3 [13.5, 22.4] | 32.7 [27, 37.5] | 38 [30.4, 43.6] | 31.1 [22, 37.9] | **120.1 [92.9, 141.4]** |
| Texas | 1.7 [0, 3.5] | 9.6 [7.5, 11.7] | 10.4 [7.9, 12.6] | 15.4 [12.6, 18] | **37.2 [27.9, 45.9]** |
| Virginia | 12 [8.8, 14.8] | 20.4 [16.8, 23.7] | 21.8 [17.5, 25.5] | 24.9 [19.8, 29] | **79.2 [63.0, 93.0]** |
| Washington | 9.5 [4.1, 14.2] | 35.8 [29.9, 40.4] | 50 [43.8, 55] | 84.5 [77.8, 89.4] | **179.8 [155.6, 199]** |
| Wisconsin | 10.7 [1, 19] | 37.5 [26.1, 47] | 40.2 [27.3, 50.9] | 42.7 [27.3, 54.6] | **131.1 [81.6, 171.6]** |

**Table 8.** Annual Excess Drug Overdose Death Rates Among Hispanic Populations, by State, 2020–2023.

| State | Rate 2020 [LB, UB] | Rate 2021 [LB, UB] | Rate 2022 [LB, UB] | Rate 2023 [LB, UB] | Total [LB, UB] |
| --- | --- | --- | --- | --- | --- |
| Arizona | 6.3 [2.7, 9.5] | 3.8 [0, 8.3] | 0 [0, 0.2] | 0 [0, 0] | 10 [2.7, 18] |
| California | 5.7 [4.7, 6.5] | 7.2 [5.9, 8.2] | 6.8 [5, 8.2] | 6.1 [3.7, 7.9] | 25.7 [19.3, 30.8] |
| Colorado | 9.2 [5.4, 12.8] | 16.9 [12.2, 20.9] | 14.4 [9.1, 19] | 14.7 [8.4, 20.1] | 55.1 [35.1, 72.8] |
| Connecticut | 2.3 [0, 8.5] | 5.2 [0, 12.7] | 8.2 [0, 17.5] | 0 [0, 7.7] | 15.7 [0, 46.3] |
| Florida | 3.3 [2, 4.5] | 5.5 [4.1, 6.9] | 5 [3.2, 6.5] | 3.5 [1.7, 5.3] | 17.3 [11, 23.2] |
| Illinois | 6.2 [4.1, 8.2] | 5.9 [3.5, 8.2] | 6.2 [3.4, 8.7] | 4.3 [1.2, 7.2] | 22.6 [12.2, 32.3] |
| Indiana | 2 [0, 6.2] | 8.2 [1, 13.3] | 3.8 [0, 10] | 8.4 [0, 15.5] | 22.3 [1, 45] |
| Massachusetts | 0 [0, 5] | 0.8 [0, 7] | 4.8 [0, 11.9] | 1.5 [0, 9.3] | 7.1 [0, 33.3] |
| Michigan | 3.7 [0, 8.1] | 11.3 [5.5, 15.9] | 8.7 [2.6, 13.8] | 3 [0, 8.3] | 26.6 [8.1, 46.1] |
| Nevada | 6.2 [3.4, 8.7] | 8.7 [5.3, 11.4] | 8.4 [4.4, 11.2] | 13.5 [8.9, 16.5] | 36.8 [22, 47.8] |
| New Jersey | 0 [0, 0.4] | 0 [0, 1.6] | 0 [0, 1.1] | 0 [0, 0] | 0 [0, 3.1] |
| New Mexico | 11.5 [6.7, 15.8] | 29 [23.7, 33.6] | 22.7 [16.5, 27.6] | 22.4 [15.9, 27.7] | 85.5 [62.8, 104.7] |
| New York | 3.6 [1.7, 5.6] | 11.3 [9.1, 13.5] | 13.7 [11.3, 16.3] | 13.5 [10.7, 16.3] | 42.2 [32.7, 51.8] |
| North Carolina | 3.6 [1.1, 5.5] | 8.6 [5.4, 10.7] | 11.9 [8.1, 14.4] | 12.9 [8.5, 15.7] | 37 [23.1, 46.3] |
| Ohio | 7.8 [1.6, 13.1] | 6.8 [0, 12.9] | 9.2 [1, 15.8] | 9.2 [0.1, 16.6] | 33 [2.8, 58.4] |
| Pennsylvania | 0 [0, 0] | 0.5 [0, 7] | 0 [0, 0] | 0 [0, 5.3] | 0.5 [0, 12.3] |
| Texas | 0.8 [0.1, 1.4] | 4.6 [3.8, 5.3] | 4.1 [3.3, 4.9] | 6.5 [5.5, 7.3] | 15.9 [12.7, 18.9] |
| Utah | 1.3 [0, 4.7] | 5.7 [0.4, 9.5] | 1.3 [0, 5.7] | 4.5 [0, 9.3] | 12.7 [0.4, 29.3] |
| Washington | 4 [1.7, 6.1] | 10.7 [8.2, 13.1] | 16 [13.1, 18.5] | 25.6 [22.5, 28.1] | 56.3 [45.4, 65.9] |
| Wisconsin | 6.1 [0.5, 10.7] | 9.6 [2.8, 14.8] | 15.7 [7.5, 21.4] | 17.9 [8, 24] | 49.3 [18.8, 70.8] |

**Table 9.** Annual Excess Drug Overdose Death Rates Among Asian Populations, by State, 2020–2023.

| State | Rate 2020 [LB, UB] | Rate 2021 [LB, UB] | Rate 2022 [LB, UB] | Rate 2023 [LB, UB] | Total [LB, UB] |
| --- | --- | --- | --- | --- | --- |
| California | 1.7 [1, 2.3] | 1.8 [1.1, 2.5] | 3.5 [2.8, 4.2] | 3.3 [2.4, 4] | 10.3 [7.3, 12.9] |
| Hawaii | 0 [0, 0] | 0 [0, 0] | 0 [0, 0] | 0 [0, 0] | 0 [0, 0] |
| New York | 0.4 [0, 1.3] | 1.3 [0, 2.3] | 2 [0.6, 3.1] | 0.8 [0, 1.9] | 4.5 [0.6, 8.6] |

**Table 10.**
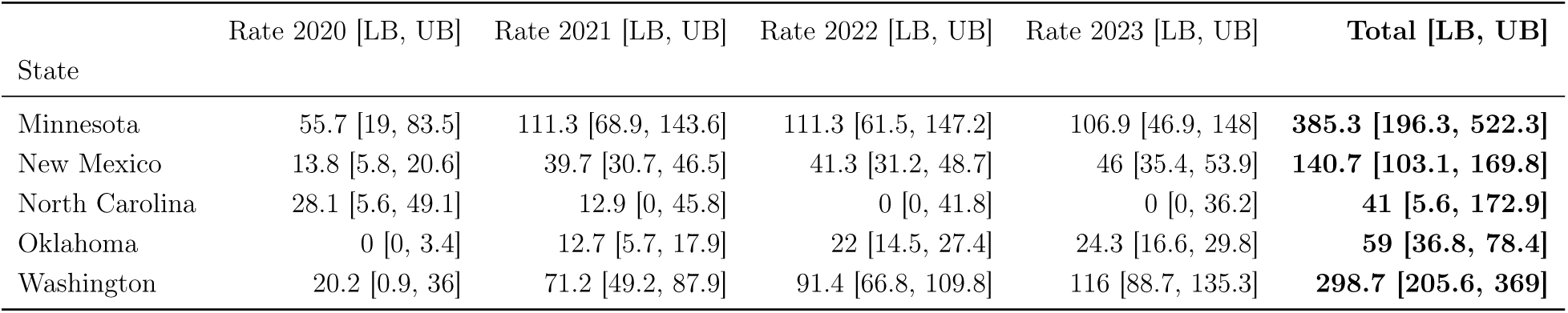
Annual Excess Drug Overdose Death Rates Among Native Populations, by State, 2020–2023.

| State | Rate 2020 [LB, UB] | Rate 2021 [LB, UB] | Rate 2022 [LB, UB] | Rate 2023 [LB, UB] | Total [LB, UB] |
| --- | --- | --- | --- | --- | --- |
| Minnesota | 55.7 [19, 83.5] | 111.3 [68.9, 143.6] | 111.3 [61.5, 147.2] | 106.9 [46.9, 148] | 385.3 [196.3, 522.3] |
| New Mexico | 13.8 [5.8, 20.6] | 39.7 [30.7, 46.5] | 41.3 [31.2, 48.7] | 46 [35.4, 53.9] | 140.7 [103.1, 169.8] |
| North Carolina | 28.1 [5.6, 49.1] | 12.9 [0, 45.8] | 0 [0, 41.8] | 0 [0, 36.2] | 41 [5.6, 172.9] |
| Oklahoma | 0 [0, 3.4] | 12.7 [5.7, 17.9] | 22 [14.5, 27.4] | 24.3 [16.6, 29.8] | 59 [36.8, 78.4] |
| Washington | 20.2 [0.9, 36] | 71.2 [49.2, 87.9] | 91.4 [66.8, 109.8] | 116 [88.7, 135.3] | 298.7 [205.6, 369] |

When comparing excess overdose rates across White, Hispanic, and Black populations, the three-group overlap in Figure 4 (N = 15 states) shows clear separation consistent with a global Friedman test indicating significant differences when blocking by state (*p* = 0.0007). Post-hoc Wilcoxon signed-rank tests with Holm correction indicate higher excess-death rates for Black populations than for White and Hispanic populations, with no meaningful difference between White and Hispanic rates (Supplementary Figures S7–S8).

**Figure 4:**
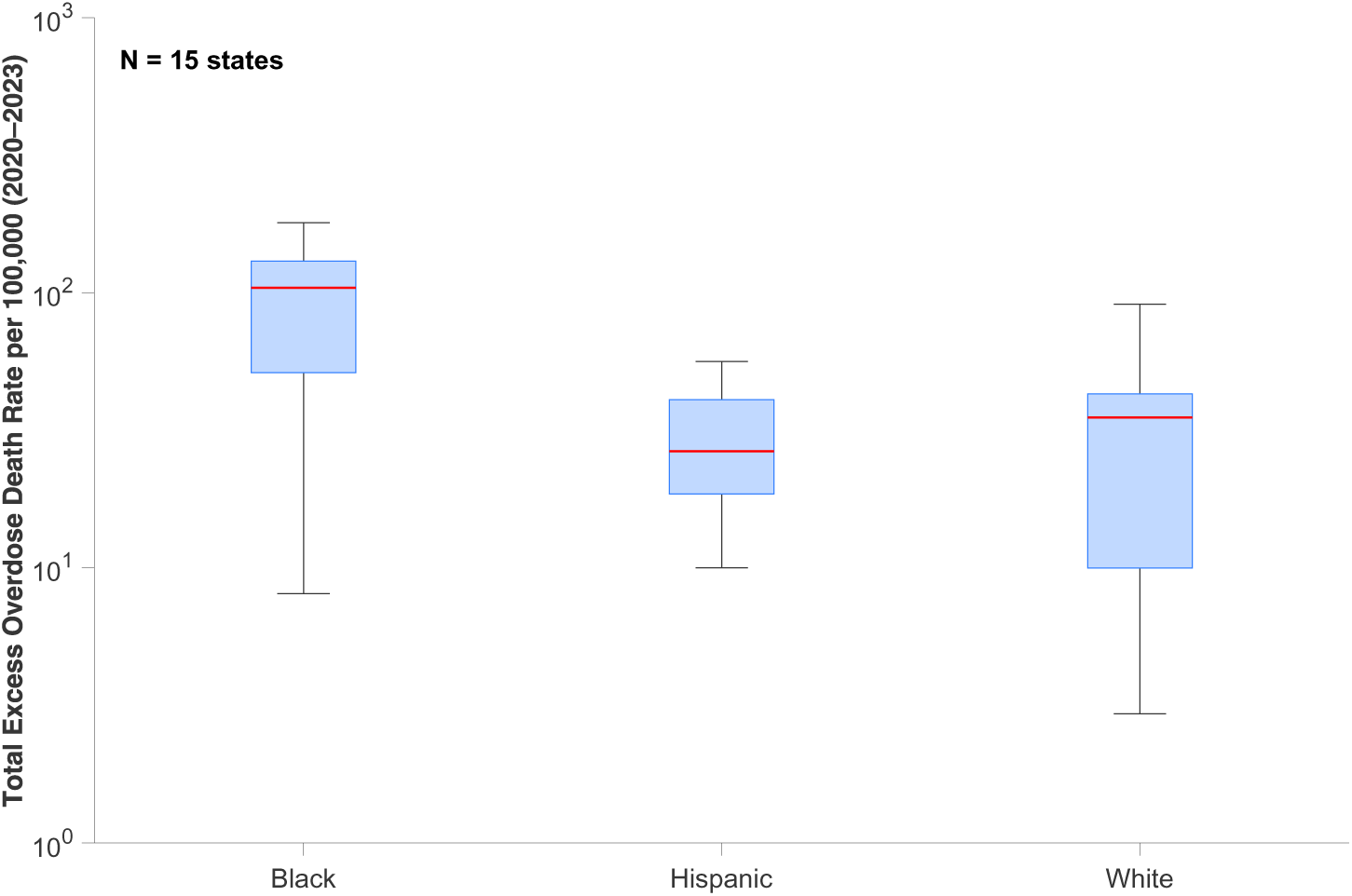
Boxplots of total excess overdose death rate per 100,000 (2020–2023) by racial group (Hispanic, Black, and White; log_10_ scale). Each box aggregates state-level excess overdose death rate estimates for states that report data for all three racial groups (N = 15 states). Red lines denote median excess-death rates; whiskers extend to 1.5×IQR. Black populations exhibit higher median excess-death rates than Hispanic and White populations.

**Figure 5:**
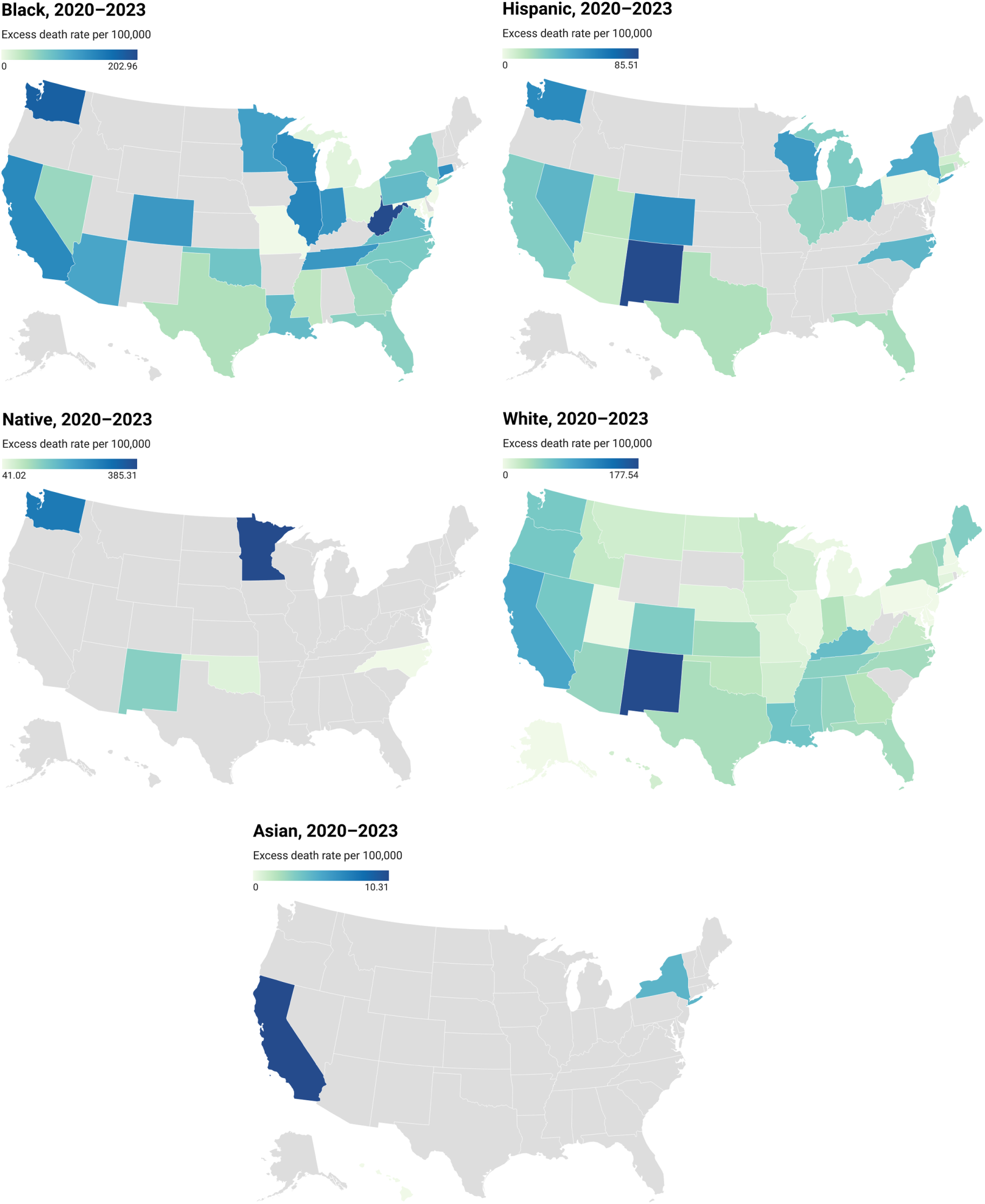
Estimated state- and race-specific excess overdose mortality rates (per 100,000), 2020–2023. Rates were obtained with a Generalized Growth Model (GGM) calibrated to 2014–2019 overdose mortality and projected through 2020–2023. Panels show Black, Hispanic, American Indian/Alaska Native, White, and Asian populations. Darker shading indicates higher excess mortality; gray denotes no available data.

## 5. Relationship Between Excess Overdose Rate and State-Level Predictors

We quantified bivariate associations (Spearman’s *ρ*) between six state-level predic-tors—alcohol use, smoking, labor force participation, poverty rate, binge-drinking prevalence, and uninsured—and excess overdose death rates (2020–2023), stratified by race/ethnicity (Supplementary Figures S9–S11).

For the White population, higher poverty was positively associated with excess overdose mortality, whereas higher labor force participation and higher alcohol use were negatively associated with excess (Supplementary Figure S9). Smoking, binge drinking, and uninsured prevalence showed weaker or non-significant associations. By contrast, in the Black and Hispanic populations, most predictors did not exhibit strong or statistically significant correlations with excess overdose mortality (Supplementary Figures S10–S11). One notable exception was a negative association between uninsured prevalence and excess overdose mortality among Black residents. Because these results are ecological and based on simple correlations, they should not be interpreted causally.

## 6. Discussion

This study provides, to our knowledge, the first state-level, race/ethnicity-stratified estimates of excess drug overdose mortality through 2023 and introduces population-scaling methods to quantify how overdose burden concentrates across population sizes. By combining excess-mortality estimation with distributional comparisons and population-scaling analysis, we characterize both the magnitude and structural distribution of overdose risk across racial/ethnic groups during the pandemic years. We find substantial excess overdose mortality from 2020–2023 in most states, with marked racial/ethnic disparities and pronounced geographic variation.

From 2020–2023, the United States experienced marked excess overdose mortality with clear racial/ethnic stratification. Absolute excess counts were largest for White populations, peaking around 2021 and declining thereafter in several states, while Black and Hispanic communities showed later but sharp increases in excess deaths per capita. These temporal and racialized patterns are consistent with national surveillance documenting that the early pandemic amplified pre-existing inequities—Black overdose mortality rose fastest from 2019 to 2020 and, in some settings, surpassed that of Whites Friedman and Hansen (2022); Kariisa (2022); Hedegaard et al. (2021).

Geographically, excess mortality was highly heterogeneous. States in Appalachia, parts of the Midwest, and the West Coast repeatedly appeared as hot spots, and race-specific maps revealed distinct regional risk profiles. Such regional clustering likely reflects shared structural conditions, variation in treatment capacity, and region-specific drug-supply dynamics, echoing earlier work on county-level clustering and spillover risk in high-burden corridors Wilt et al. (2019); Choi et al. (2022). During the study window, illicitly manufactured fentanyl continued to diffuse nationally and was increasingly detected with other substances; polysubstance combinations (e.g., stimulants with opioids) and, more recently, xylazine adulteration heightened fatality risk and varied by region Ciccarone (2021); Kariisa (2023). These supply-side shifts, interacting with structural inequities in prevention and treatment access, likely contributed to the later but intense per capita surges among Black and Hispanic populations despite Whites showing the largest absolute counts Friedman and Hansen (2022).

A key contribution of our study is the population-scaling analysis. In log–log regressions, overdose burden did not increase uniformly with population size. Among Black populations, total overdose deaths scaled sublinearly with state population, and excess deaths were consistent with sublinearity, indicating disproportionately high per capita burden in smaller Black populations even where absolute counts were lower. For White and Hispanic populations, excess and total deaths scaled approximately proportionally with population size, suggesting that the absolute burden in these groups tends to rise in step with population. At the aggregate (all-race) level, point estimates exceeded proportionality, but evidence for superlinear scaling was only suggestive. These scaling patterns imply that fixed per capita allocation formulas may systematically under-resource smaller Black populations experiencing disproportionate burden, while large White and Hispanic populations require capacity scaled to the absolute volume of need. Prevention, treatment, and harm-reduction strategies should therefore be tailored to distinct scaling patterns rather than relying on uniform, population-proportional formulas.

We also examined state-level correlates of excess overdose death rates. Among Whites, higher poverty and lower labor force participation were positively associated with excess mortality rates, whereas higher labor force participation and higher alcohol use were inversely associated (Supplementary Figure S9). These ecological associations align with prior evidence that worsening local macroeconomic conditions track higher overdose risk over time, especially for White adults Hollingsworth et al. (2017); Rudolph et al. (2020). For Black and Hispanic populations in our study, associations with the same predictors were weaker or mixed, echoing evidence that economic indicators do not map uniformly onto overdose risk across racial/ethnic groups and that additional structural factors and a changing drug supply likely mediate risk Rudolph et al. (2020); Barnett et al. (2023); Dowell (2024).

The COVID-19 pandemic introduced risk amplifiers—social isolation, economic hardship, disruptions to treatment and harm reduction, and a more lethal drug supply—that hit minority communities especially hard. Our findings underscore the need for equity-focused overdose responses: improving race-specific, timely surveillance (including excess-death monitoring), expanding culturally tailored harm reduction and treatment in Black and Hispanic neighborhoods, and directing resources both to small, high-risk Black communities and to large Hispanic centers. In regions where fentanyl and stimulant co-involvement are driving large absolute burdens (e.g., many West Coast and Northeast jurisdictions), interventions must address polysubstance risk and high-volume need. In states with smaller Black populations but disproportionately high per capita excess mortality, focused community-level investment is warranted even where total case counts are modest.

Although our analyses are stratified by race and ethnicity, important within-group heterogeneity remains. Prior work shows that overdose risk is highly concentrated in specific subgroups: among adults aged 55 years and older, non-Hispanic Black men have the highest and fastest-growing opioid overdose mortality, with rates roughly four times higher than the overall rate in that age group by 2019 and a sharp acceleration as fentanyl entered the drug supply Mason et al. (2022). Recent national surveillance likewise indicates disproportionately high, pandemic-era increases among older Black men, linked to both structural disadvantage and the spread of illicitly manufactured fentanyl Volkow (2024). Because several states had sparse or suppressed counts, we could not disaggregate by age or sex, limiting our ability to assess this critical dimension of within-group variability. These patterns suggest that harm-reduction and treatment responses must be targeted not only by geography and race/ethnicity but also by high-risk demographic subgroups—particularly older Black men. This study has limitations. First, we relied on state-level race-stratified mortality data, and several states with incomplete reporting—particularly for American Indian/Alaska Native and Asian populations—were excluded, likely leading to underestimation of excess mortality in those groups. Second, we used 2020 population denominators and potentially imperfect death certification, so rate estimates may be biased, and excess mortality may reflect both misclassified drug deaths and indirect mortality related to pandemic disruptions. Third, our ecological correlation analyses are susceptible to ecological fallacy: aggregate associations can obscure within-state heterogeneity and conflate contextual factors (e.g., economic structure, service availability, or reporting completeness) with individual behavioral risks. Finally, when summarizing across years, we did not explicitly model serial dependence in mortality (i.e., temporal correlation between years), which may lead to understated uncertainty intervals.

Future work should disaggregate excess mortality by drug class, age, and sex/gender to characterize substance-specific and demographic patterns and assess how social determinants (e.g., unemployment, poverty, housing instability, and healthcare access) and service coverage contribute to interstate and within-state variation. Efforts should also advance data equity by partnering with tribal, territorial, and local agencies to fill persistent gaps for American Indian/Alaska Native and other underrepresented populations and to compare alternative baselines and modeling frameworks using prospective validation and sensitivity analyses. Finally, integrating quantitative estimates with qualitative, community-engaged inquiry will help clarify mechanisms and inform structurally responsive intervention strategies.

In summary, this study documents uneven excess overdose mortality across U.S. states during 2020–2023, showing how pre-existing structural inequities and a shifting drug supply shaped pandemic-era outcomes. Beyond the overall burden, we find heterogeneous population scaling and race-specific relationships to state-level predictors. These patterns argue for adaptive, equity-focused responses that pair ongoing, race-disaggregated monitoring with allocation rules and interventions calibrated to both scaling heterogeneity and group-specific risk structures. Future work should age-standardize rates and evaluate alternative baselines to ensure robustness while guiding sustainable strategies that improve access to harm reduction and treatment without reinforcing disparities.

## Data Availability

All data produced in the present study are available upon reasonable request to the authors.

https://vizhub.healthdata.org/gbd-results/

https://www.kff.org/state-health-policy-data/state-indicator/people-0-64-uninsured-rate-by-age/?currentTimeframe=0&sortModel=%7B%22colId%22:%22Location%22,%22sort%22:%22asc%22%7D

https://www.cdc.gov/alcohol/excessive-drinking-data/index.html

https://fred.stlouisfed.org/graph/?graph_id=225188

https://www.census.gov/programs-surveys/saipe/about/faq.html

## Supplementary Methods

### S1. Data Sources and State-Level Predictors

The dataset used in this study comprises annual, state-level counts of drug overdose deaths in the United States, stratified by race/ethnicity and year for the period 2014 through 2023. Mortality data were obtained from public health surveillance systems managed by the Centers for Disease Control and Prevention (CDC), specifically from the CDC WONDER database Multiple Cause of Death Data on CDC WONDER. To ensure consistent coverage and maximize completeness across the study period, we combined two overlapping data sources: the Current Final MCOD Data for 1999–2020 and the Current Final MCOD Data for 2018–2023. Because these files overlap in reporting 2018–2020, we retained counts from the 1999–2020 series for overlapping years to preserve internal consistency in coding conventions, data processing, and population estimates.

Drug overdose deaths were identified using the International Classification of Diseases, 10th Revision (ICD-10) codes. The codes included for the underlying cause of death were X40–X44 (unintentional), X60–X64 (suicide), X85 (homicide), and Y10–Y14 (undetermined), which are consistent with definitions used in prior studies Billock et al. (2023). These categories capture all injury-related drug poisonings regardless of intent, enabling comparability with CDC and peer-reviewed overdose surveillance methodologies. Records associated with other ICD-10 codes were excluded.

Population denominators were derived from the CDC Bridged-Race Population Estimates, which provide annual population counts by year, state, race (four categories), ethnicity, sex, and age. We used the 2020 population estimates as a fixed reference denominator across all study years to compute rates Bridged-Race Population Estimates. This provided a consistent reference point across states and racial/ethnic groups. Supplementary Figure S1 illustrates the racial/ethnic composition of each state, highlighting the relative population sizes that underlie the per capita overdose mortality estimates presented in this study.

We compiled six state-level socioeconomic and health-behavior indicators from Global Burden of Disease (GBD), CDC, KFF, BLS/FRED, and the Census SAIPE program. All indicators represent the total state population (not race-specific strata):

- **Alcohol use (GBD)** is defined as the prevalence of current drinkers who consumed ≥1 alcoholic beverage in the past 12 months Institute for Health Metrics and Evaluation (IHME) (2025).
- **Smoking (GBD)** is defined as the prevalence of any current use of smoked tobacco (daily or occasional) Institute for Health Metrics and Evaluation (IHME) (2025).
- **Binge drinking (CDC)** is the percentage of adults (≥18 years) who report consuming ≥5 drinks (men) or ≥4 drinks (women) on one occasion during the past 30 days, based on BRFSS data Centers for Disease Control and Prevention (2024).
- **Uninsured rate (KFF)** is the share of the total population without health insurance, including those with Indian Health Service–only coverage KFF State Health Facts (2025).
- **Labor force participation (BLS/FRED)** is the number of people in the labor force as a percentage of the civilian noninstitutional population—that is, those working or actively seeking work Federal Reserve Bank of St. Louis (2025).
- **Poverty rate (Census SAIPE)** is the percentage of people living below the official federal poverty thresholds, using SAIPE’s annual model-based state estimates U.S. Census Bureau (2025).

### S2. Data Preparation and State Inclusion

The primary racial and ethnic categories considered in the analysis include White, Black, Hispanic, Asian, and Native American populations. Each observation in the dataset includes the following variables: state, year (2014–2023), racial/ethnic group, and the number of reported drug overdose deaths.

For each racial/ethnic group, data were available from multiple U.S. states, although reporting completeness varied across jurisdictions and years. To support reliable intergroup comparisons, analyses were restricted to state–race combinations that provided complete data over the study period. For example, California reported complete data for all five racial/ethnic groups across all years and was fully included. In contrast, New York was missing data for the Native American population in some years, so only the four fully reported racial groups were analyzed for that state. States such as South Dakota, Rhode Island, and Wyoming, which lacked complete data for all racial groups across all years, were excluded entirely. The District of Columbia, although not a state, was included due to its complete reporting. In total, the final analysis included 47 states and the District of Columbia.

Some states had missing data for specific groups, particularly for smaller populations such as Asian and Native American individuals. The final dataset allows for both disaggregated and aggregated analyses of overdose mortality. While the primary unit of analysis is the state–year–race combination, additional aggregations were performed for visualizations and summary statistics in the Results section. In addition to comparing excess overdose deaths, we also analyzed the overall distribution of excess overdose death rates across the racial groups included in each comparison (Figure 4 in the main text, Supplementary Figures S7–S8). Rather than evaluating state-by-state patterns, we aggregated data from all states that had complete records for the racial groups being compared. This allowed us to present a clear view of how excess death rates are distributed within each group, ensuring that the comparisons were based on consistent and reliable data.

### S3. Generalized Growth Model (GGM)

The GGM is a two-parameter growth model that flexibly captures dynamics ranging from sub-exponential to exponential growth Chowell et al. (2019); Viboud et al. (2016). This flexibility makes the GGM well suited for describing the nonlinear pre-2020 upward trend in overdose deaths. The model is defined by the differential equation

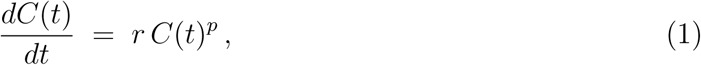

where *C*(*t*) is the cumulative number of overdose deaths at time *t*, *r >* 0 is the growth rate, and 0 ≤ *p* ≤ 1 is the *growth exponent* that controls curvature. For *p* ≠ 1, Equation (1) admits the closed-form solution

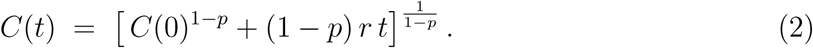

For *p* = 1, the solution reduces to the exponential form *C*(*t*) = *C*(0) *e^rt^*.

In our application, for each state–race stratum, we set the initial condition at the first calibration year, *C*(0) = *D*_2014_, where *D*_2014_ denotes the observed deaths in 2014. We then estimate the parameters (*r, p*) by maximum likelihood under a Poisson model for the annual increments *Y_t_* (2015–2019), with mean *µ_t_* = *C*(*t*; *r, p*) − *C*(*t* − 1; *r, p*), where *C*(*t*) is the cumulative count implied by the GGM. We project to 2020–2023 by evaluating Equation (2) at *t* ∈ {2020, 2021, 2022, 2023}, with corresponding projected annual deaths *Ŷ_t_* = *Ĉ*(*t*) − *Ĉ*(*t* − 1). We implemented this using the SubEpiPredict toolbox Chowell et al. (2024).

### S4. Simple Linear Regression (SLR)

For comparison, we also modeled the mortality trend with a simple linear regression on year. The SLR assumes overdose deaths increase (or decrease) at a constant absolute rate over time. For a given dataset, we fit an ordinary least squares linear model

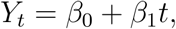

where *Y_t_* is the number of overdose deaths in year *t*, *β*_0_ is the intercept, and *β*_1_ is the annual slope. The model was fitted to the 2014–2019 data points to obtain the best-fit line, which was then extrapolated to predict annual deaths in 2020–2023. We implemented SLR fits and forecasts using the StatModPredict R-Shiny toolbox Bleichrodt et al. (2024). The SLR provides a baseline trend assuming a steady linear growth rate, against which the more flexible GGM can be compared.

### S5. Correlation and Calibration Analyses

We assessed the correlation between the two models’ predictions across state and racial/ethnic groups with available data. We gathered the annual predicted values from 2020–2023 for both the GGM and the SLR and calculated Spearman’s rank correlation coefficient *ρ* between the two series. In our analysis, we observed a strong positive *ρ* = 0.98 (Supplementary Figure S2), indicating that, when aggregated across all states and races, the GGM and SLR forecasts exhibited strong consistency in projected deaths.

In addition to the correlation assessment, we evaluated the calibration performance of both models by comparing their predictions for 2014–2019 against the observed overdose deaths in those years. Let *Y* (*t*) denote the observed death count in year *t* and *Y*^^^(*t*) the model’s prediction. For each model and each state–race stratum, we computed the Mean Absolute Error (MAE) and Mean Squared Error (MSE) over the six calibration years (2014–2019) as follows:

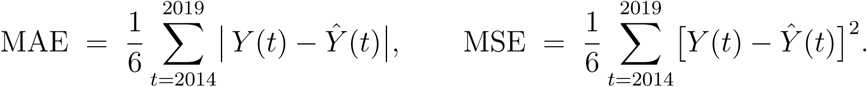

MAE measures the average magnitude of the errors in absolute terms, while MSE penalizes larger deviations by squaring the residuals Hyndman and Koehler (2006). Lower values indicate better agreement between predicted and observed values. Across all state–race strata with available data, we computed the MAE and MSE for each model’s 2014–2019 predictions and then summarized performance by taking the mean across those strata. The GGM yielded a lower average MAE and MSE than the SLR (Supplementary Figure S2), indicating superior calibration of the nonlinear growth model to the pre-2020 trajectory.

### S6. Excess-Death and Rate Calculations

We focused on two parsimonious growth models for their interpretability and suitability to short time series. Based on calibration and correlation results, we selected the GGM as the preferred baseline for estimating excess overdose deaths. Excess overdose deaths attributable to unforeseen factors (e.g., the COVID-19 pandemic and its societal impacts) were calculated as

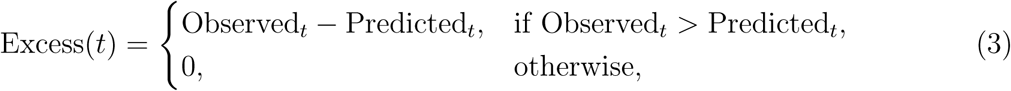

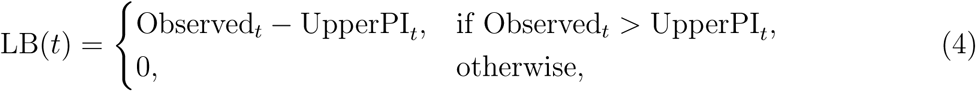

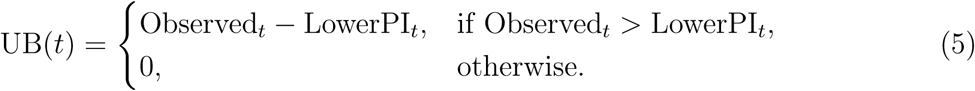

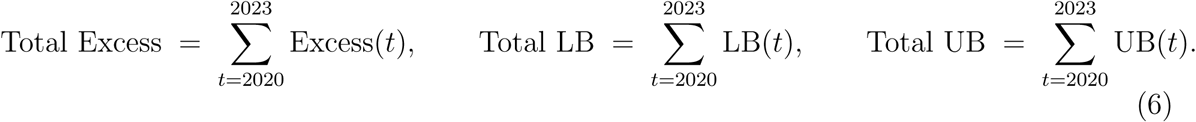

Let Pop_2020_ denote the population size in 2020 (fixed denominator). Annual and cumulative rates were computed as

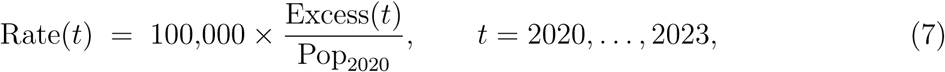

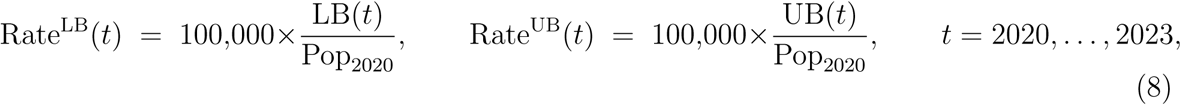

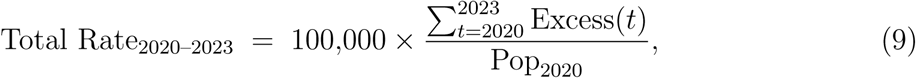

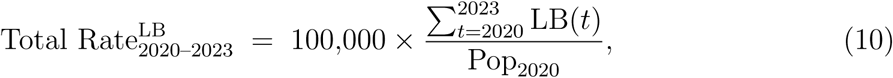

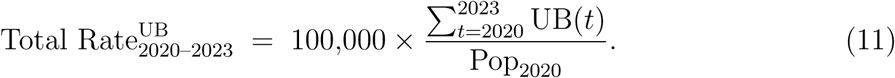

We use a standard approach to estimate excess deaths Weinberger et al. (2020). The model estimates how many deaths would have occurred if past trends had continued (the counterfactual). Excess deaths are the difference between observed and expected values.

This baseline versus observed method has been previously used to assess the impact of large-scale crises Karami et al. (2025); Zhao et al. (2024); Kontis et al. (2020). Although prediction intervals are not additive across years because the uncertainty of the total depends on cross-year covariance, as a transparent, conservative summary, we report a sum-of-bounds interval obtained by adding annual lower and upper bounds. Given that pandemic-year shocks were not perfectly synchronized, this procedure likely overstates uncertainty relative to a jointly estimated multi-year interval.

## Results

### S1. White Populations

Across the states with complete data, the cumulative excess deaths in White populations over 2020–2023 were greater than those of other groups (Supplementary Figures S3–S5). For example, Supplementary Figure S3 shows that the total excess overdose mortality among Whites exceeded that of Blacks in most of the comparable states except Connecticut, Georgia, and Illinois. Supplementary Figure S4 shows a similar pattern of higher White excess deaths compared to Hispanics.

In the combined three-group comparison (Supplementary Figure S5), White populations had the largest excess mortality burden, followed by Black and then Hispanic populations. These differences were reflected in state-level estimates: in California, the White population saw an estimated 3,743 excess overdose deaths in 2023 alone (95% CI: 3,495–4,012), nearly matching the peak value observed in 2021 (≈ 3,755 excess deaths) (Table 1 in the main text). Other large states also showed substantial excess mortality among White residents. Florida’s White population reached approximately 1,635 excess deaths in 2021 (95% CI: 1,425–1,889), followed by a decrease to about 398 in 2023 (95% CI: 147–709), suggesting partial attenuation of the early-pandemic surge. Kentucky also displayed persistently high burdens, recording 829 excess White overdose deaths in 2021 (95% CI: 738–911) and remaining elevated with 519 in 2023 (95% CI: 424–606).

### S2. Black Populations

Black populations suffered a pronounced increase in excess overdose mortality during 2020–2023, with particularly large surges in certain states. Notably, many states reported little to no excess overdose deaths among Black residents in 2020 but saw significant excess mortality in subsequent years. For example, Arizona’s Black population had essentially zero excess overdose deaths in 2020, yet by 2021 it experienced 128 excess deaths (95% CI: 97–154) and remained elevated through 2023. Across most states with available data, the total excess deaths among Black Americans from 2020–2023 were lower in absolute number than among Whites (Supplementary Figure S3).

Several states with large Black populations showed substantial and growing burdens. California recorded 1,243 excess Black overdose deaths in 2023 (95% CI: 1,180.5–1,291), contributing to a four-year total of over three thousand excess deaths in California’s Black community. Florida’s Black population peaked at 716 excess deaths in 2022 (95% CI: 659–765), and Georgia saw excess deaths climb from 436 in 2021 to 633 in 2023 (95% CI: 589–666). By 2023, Georgia’s Black excess (≈ 633) was comparable to Georgia’s White excess (≈ 632), despite a smaller Black population. A similar convergence occurred in the District of Columbia, where Black residents suffered 232 excess deaths in 2021 (95% CI: 196–264), followed by 210 in 2022 and 193 in 2023, figures that are very close to White overdose excess in D.C.

State-level excess deaths among Black populations display wide variation; on the log scale, central tendency and spread are only modestly lower than for White populations. California, Florida, and Georgia had the highest cumulative Black excess deaths (Supplementary Figure S6; Table 2 in the main text).

### S3. Hispanic Populations

Hispanic populations also endured notable excess overdose mortality from 2020 to 2023, though in most states the absolute numbers were lower than in White or Black populations. In aggregate comparisons, the total excess deaths among Hispanics were significantly below those of Whites (Supplementary Figure S4), making Hispanics the lowest of the three groups in the cumulative 2020–2023 burden illustrated in Supplementary Figure S5. State-level patterns underscore this disparity: across many states, Hispanic communities exhibit markedly lower excess-death counts than White and Black populations, and in numerous jurisdictions there was little or no measurable Hispanic surge (Supplementary Figure S6).

Nonetheless, certain states with large Hispanic populations faced substantial overdose mortality increases. California saw the greatest impact: the Hispanic population in California had approximately 1,116 excess overdose deaths in 2021 (95% CI: 920.5–1,283) and remained high with 948 by 2023 (95% CI: 575.5–1,232.5). This four-year total for California Hispanics (roughly 4,000 excess deaths) was higher than that of California’s Black population, though still only about one-third of the White excess deaths in that state. Florida’s Hispanic population experienced a peak of 319 excess deaths in 2021 (95% CI: 236–398), which declined to 203 in 2023 (95% CI: 97–302), showing the temporal pattern seen in Florida’s White population (early pandemic surge followed by improvement). In New York, excess overdose deaths among Hispanics rose to 514 by 2022 and slightly decreased in 2023 (506) (Table 3 in the main text), approaching the levels observed in New York’s White population. Many other states showed more modest excess deaths for Hispanics; for instance, Illinois had on the order of only ≈ 100 excess Hispanic overdose deaths per year by 2023, and some states had virtually none in certain years (e.g., Arizona saw no excess in 2022–2023 after a spike in 2020).

### S4. Asian Populations

Asian populations experienced the smallest excess overdose mortality impact in the study, and data were available for only three states. In states with smaller Asian communities, no significant excess deaths were detected for 2020–2023 (e.g., Hawaii reported zero excess overdose deaths among Asians in every year). In the three states with data, excess counts remained modest. California’s Asian population suffered an estimated 227 excess overdose deaths in 2022 (95% CI: 177–270), decreasing to 211 in 2023 (95% CI: 155–255), up from 107 in 2020—indicating a meaningful but limited surge. New York saw 37 excess deaths in 2022 (95% CI: 11–56) and 15 in 2023, while a third measured state showed none across all years. The state-level map for Asians (Supplementary Figure S6) is dominated by lighter shades, reflecting low or absent excess in most areas, with darker shades in California and New York. In general, Asian Americans had low excess overdose mortality during 2020–2023, further restricted by very limited data coverage.

### S5. Native American Populations

Excess overdose deaths among Native American populations showed a heterogeneous pattern across states, with a few states experiencing pronounced surges while others saw minimal impact. Nationally, the total excess deaths in this group were much lower in absolute count than those in White, Black, or Hispanic populations (consistent with the smaller population size and the fact that data were available for fewer states). Nevertheless, certain states with substantial Native American communities registered notable spikes. Washington State had the highest documented excess overdose mortality for Native populations, reaching 132 excess deaths in 2023 (95% CI: 101–154) after increasing each year from 2020 onward. New Mexico similarly showed a steady increase, from 26 excess deaths in 2020 to 87 in 2023 (95% CI: 67–102), reflecting a compounding burden in that state’s Native population. Oklahoma saw a surge as well, with excess deaths climbing to 94 by 2023 (95% CI: 64–115). In contrast, some states experienced early spikes that later declined. For example, Minnesota had 38 Native excess deaths in 2020 and about 76 in 2021–2022, then slightly lower at 73 in 2023, while North Carolina peaked at 35 in 2020 but had effectively zero excess overdose deaths by 2022–2023. The geographic distribution of the excess burden in Native communities is highlighted in Supplementary Figure S6 and Table 5 in the main text.

### S6. Excess Overdose Rates and Within-State Comparisons

When comparing excess overdose rates across White, Hispanic, and Black populations, the three-group overlap in Figure 4 in the main text (N = 15) shows clear separation consistent with a global Friedman test indicating significant differences when blocking by state (*p* = 0.0007). Post-hoc Wilcoxon signed-rank tests with Holm correction reinforce these differences: Black populations have higher rates than White (adjusted *p* = 0.0001; 23 states; Supplementary Figure S8) and Hispanic (adjusted *p* = 0.0006; 16 states), with Hodges–Lehmann estimates indicating that excess overdose rates are, on average, 45.5 per 100,000 higher in Black vs. White (95% CI: 19.7 to 69.7) and 72.5 per 100,000 higher in Black vs. Hispanic (95% CI: 22.7 to 102.3). In contrast, the White–Hispanic comparison (Supplementary Figure S7; 18 states) shows no meaningful difference (adjusted *p* = 0.97; HL difference = −3.2, 95% CI: −12.1 to 19.7).

### S7. Relationship Between Excess Overdose Rate and State-Level Predictors

We quantified bivariate associations (Spearman’s *ρ*) between six state-level predic-tors—alcohol use, smoking, labor force participation (LFP), poverty rate, binge-drinking prevalence, and uninsured—and excess overdose death rates (2020–2023), stratified by race/ethnicity (Supplementary Figures S9–S11).

For the White population (Supplementary Figure S9), three predictors were statistically significant: higher poverty was associated with higher excess overdose mortality (*ρ* ≈ 0.56, *p <* 0.01), whereas higher LFP and higher alcohol use were associated with lower excess (*ρ* ≈ −0.51, *p <* 0.01; *ρ* ≈ −0.36, *p <* 0.05, respectively). Smoking (*ρ* ≈ 0.10), binge drinking (*ρ* ≈ −0.16), and uninsured (*ρ* ≈ 0.26) showed weaker, non-significant relationships.

By contrast, in the Black (Supplementary Figure S10) and Hispanic (Supplementary Figure S11) populations, most predictors did not exhibit strong or statistically significant correlations with excess overdose mortality. For Black residents, approximate correlations were: alcohol 0.28, smoking −0.26, LFP 0.17, poverty −0.13, binge drinking 0.14 (all *p >* 0.05). For Hispanic residents, approximate correlations were: alcohol −0.12, smoking 0.16, LFP −0.23, poverty 0.28, binge drinking 0.21, uninsured −0.03 (all *p >* 0.05). However, among Black populations, the proportion uninsured was negatively correlated with excess overdose mortality (*ρ* ≈ −0.40, *p <* 0.05).

Because these results are ecological and based on simple correlations, they should not be interpreted causally.

---

**Figure S1:**
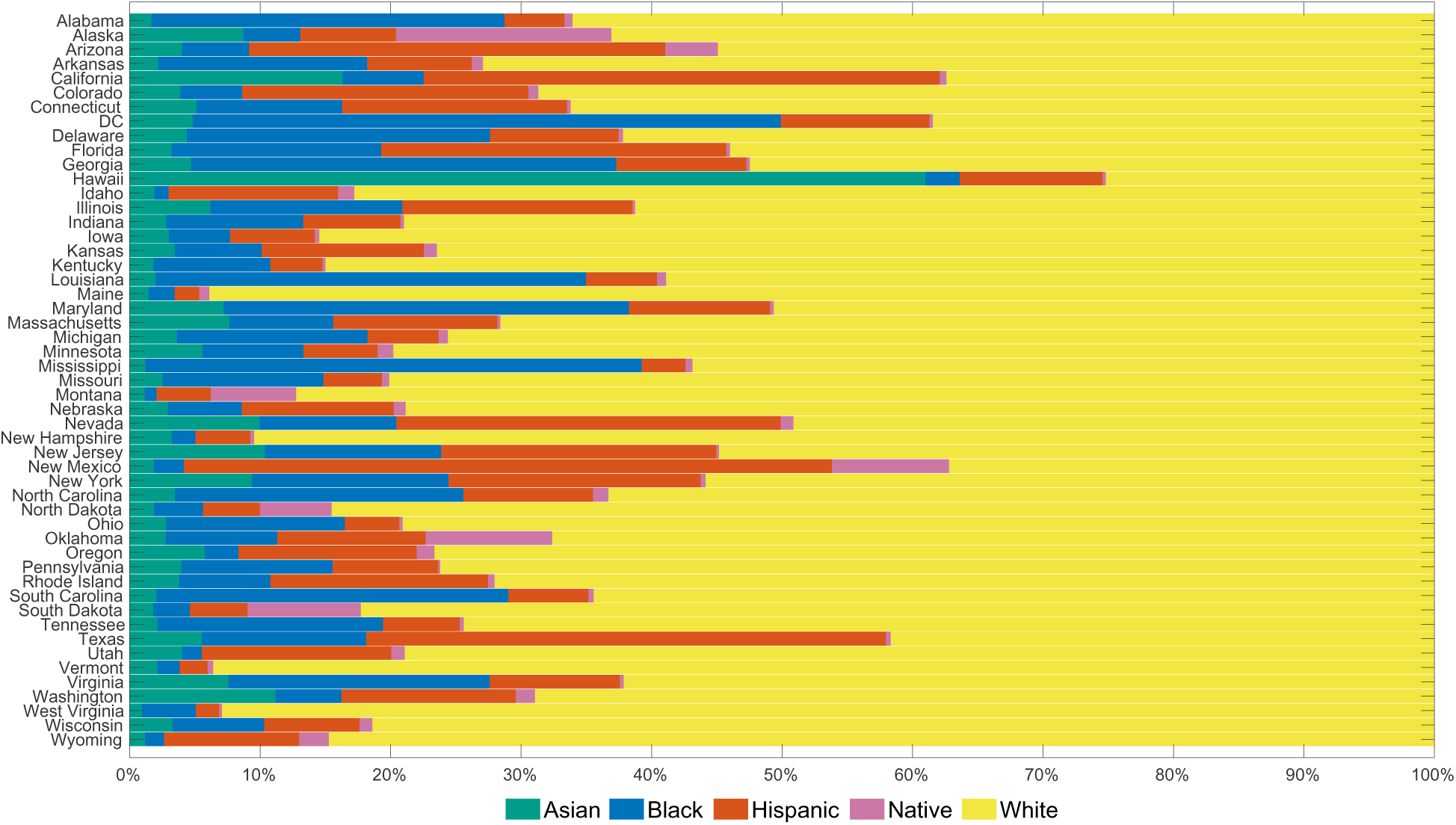
Racial/ethnic composition of state populations in 2020. Each bar represents the proportion of Hispanic, Asian, Black, Native, and White residents in a given U.S. state. This figure illustrates the demographic denominators underlying state-level excess overdose mortality estimates, providing context for interpreting differences across states and racial/ethnic groups.

**Figure S2:**
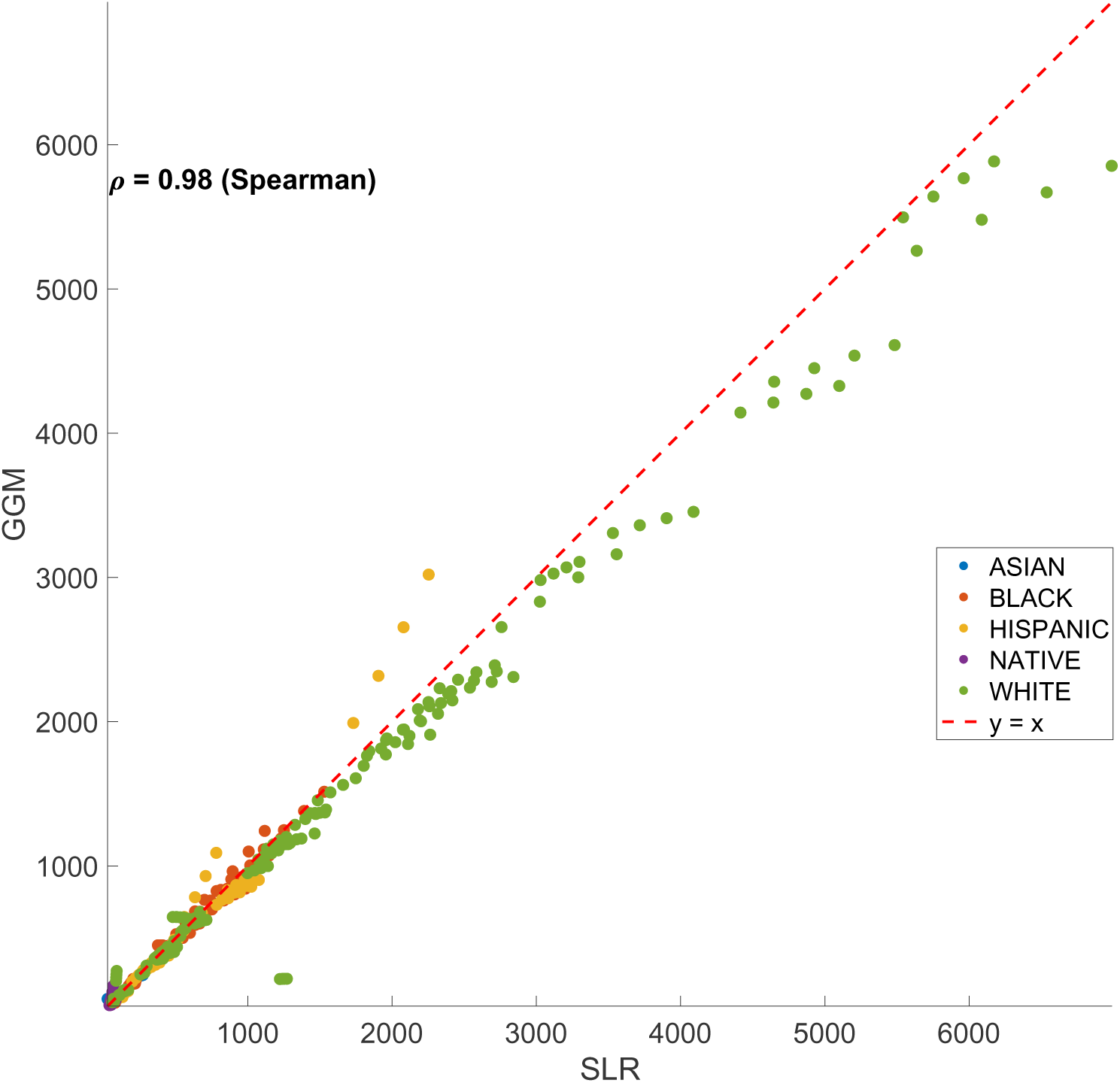
Scatter plot of predicted annual overdose deaths from the Generalized Growth Model (GGM) versus Simple Linear Regression (SLR) across all state–race strata for 2020–2023. Points are color-coded by racial group—Asian (blue), Black (orange), Hispanic (yellow), Native (purple), and White (green). The dashed red line marks the 1:1 reference, and the Spearman rank correlation coefficient (*ρ* = 0.98) indicates a very strong correlation in the relative ordering of predictions between the two models.

**Figure S3:**
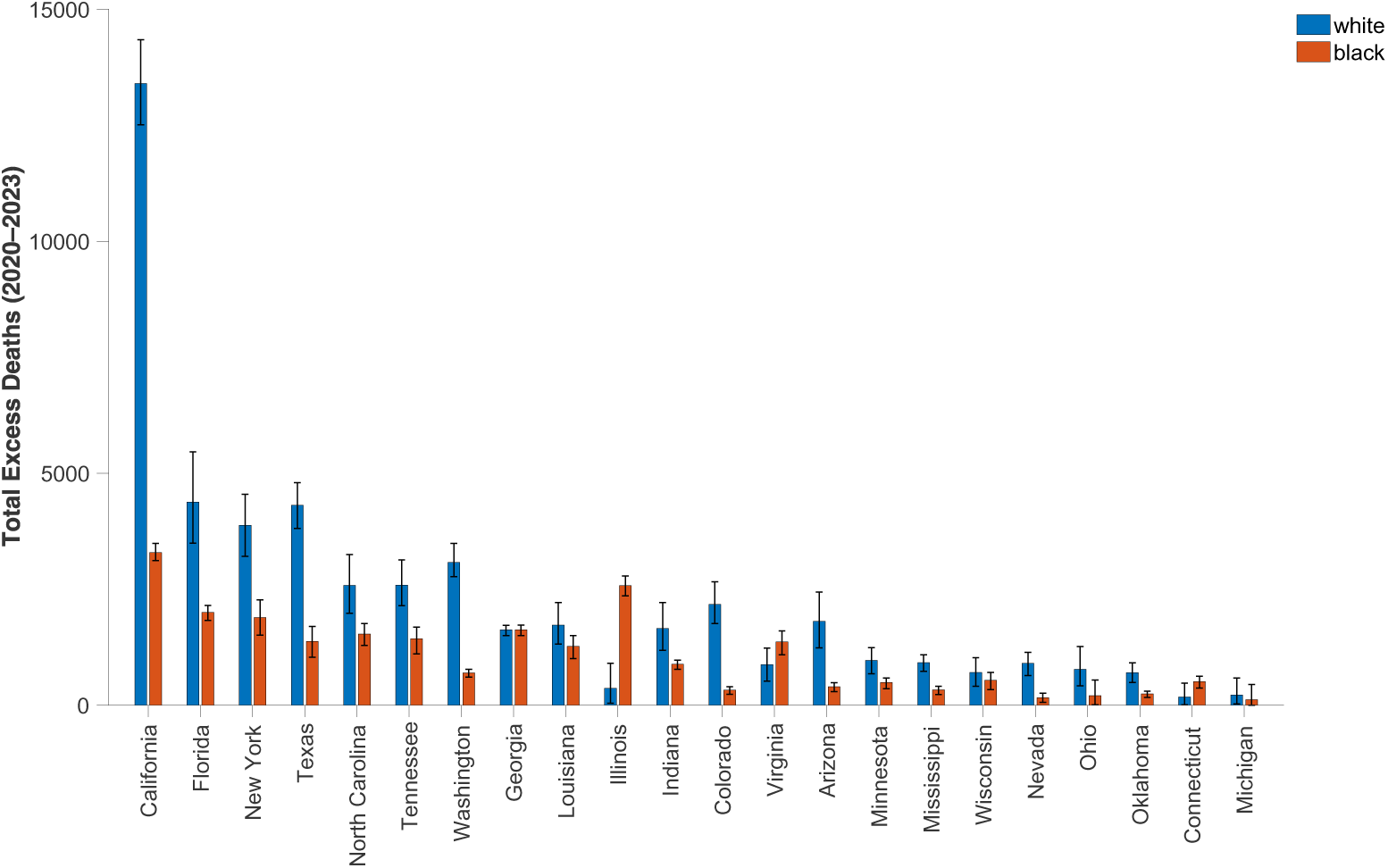
State-level total excess overdose deaths for Black (orange) and White (blue) populations over 2020–2023, with 95% PI bars. In nearly every state shown, White populations experienced larger excess death counts than Black populations, with the greatest disparities in California and Florida.

**Figure S4:**
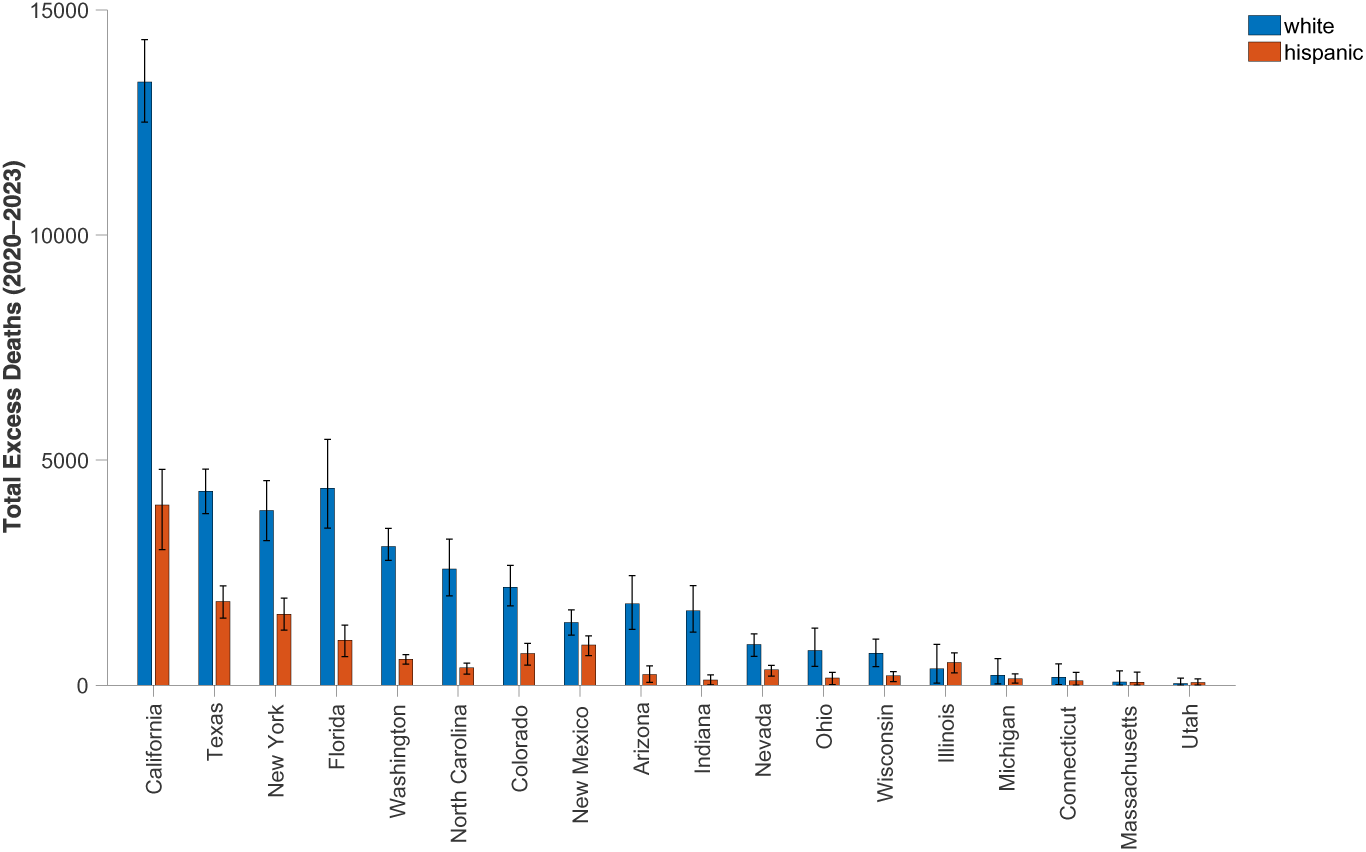
State-level total excess overdose deaths for White (blue) and Hispanic (orange) populations over 2020–2023, with 95% PI bars. White populations experienced substantially higher excess deaths in all displayed states, most dramatically in California, Texas, and Florida. Hispanic excess deaths are lower overall but show notable burdens in California, Texas, and New York. Error bars reflect uncertainty in the expected-death baseline.

**Figure S5:**
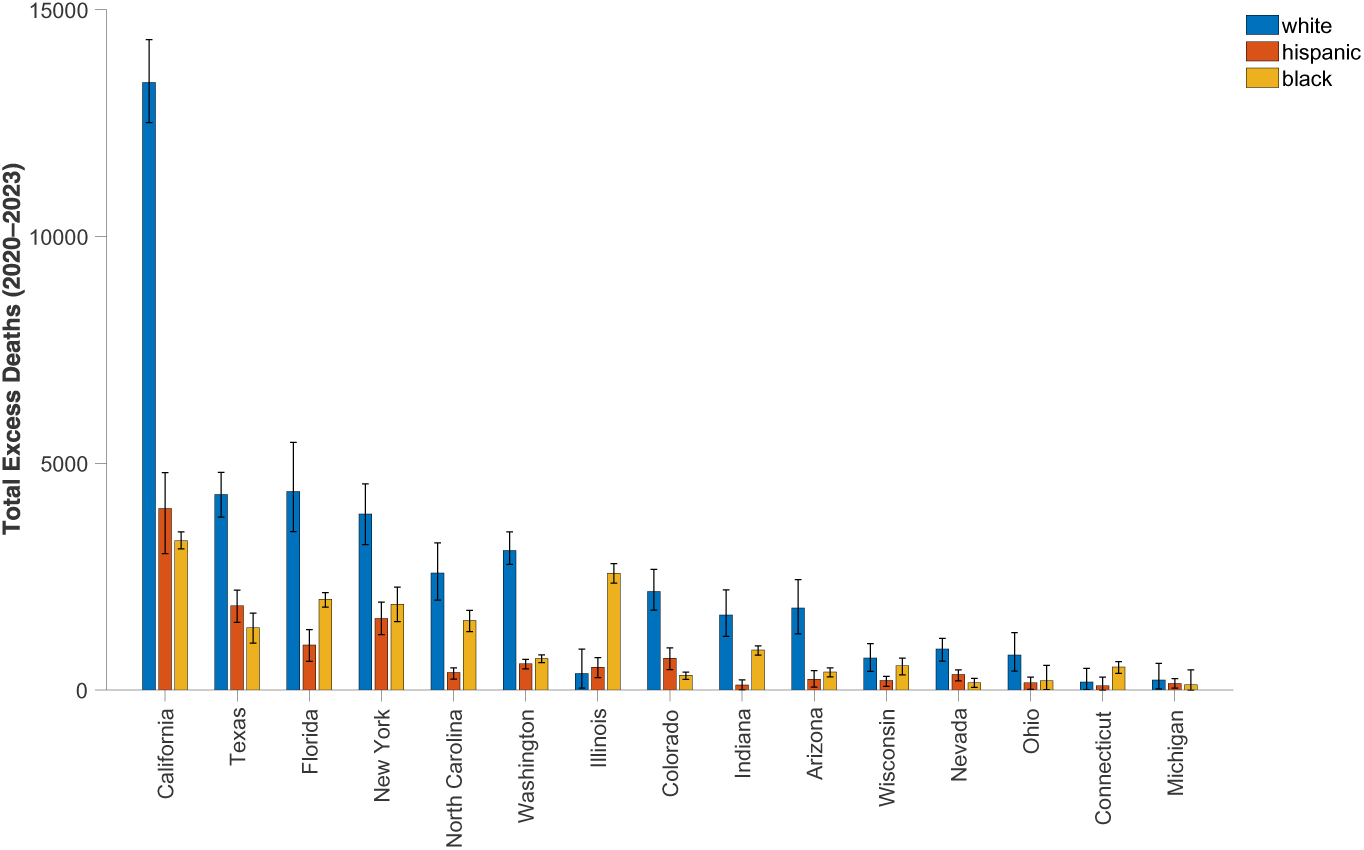
State-level total excess overdose deaths for White (blue), Black (gold), and Hispanic (orange) populations over 2020–2023, with 95% PI. White populations exhibit the highest excess death counts in almost every state, followed by Black and then Hispanic populations. The largest absolute disparities occur in California, Florida, New York, and Texas. Error bars indicate uncertainty in the expected-death baseline.

**Figure S6:**
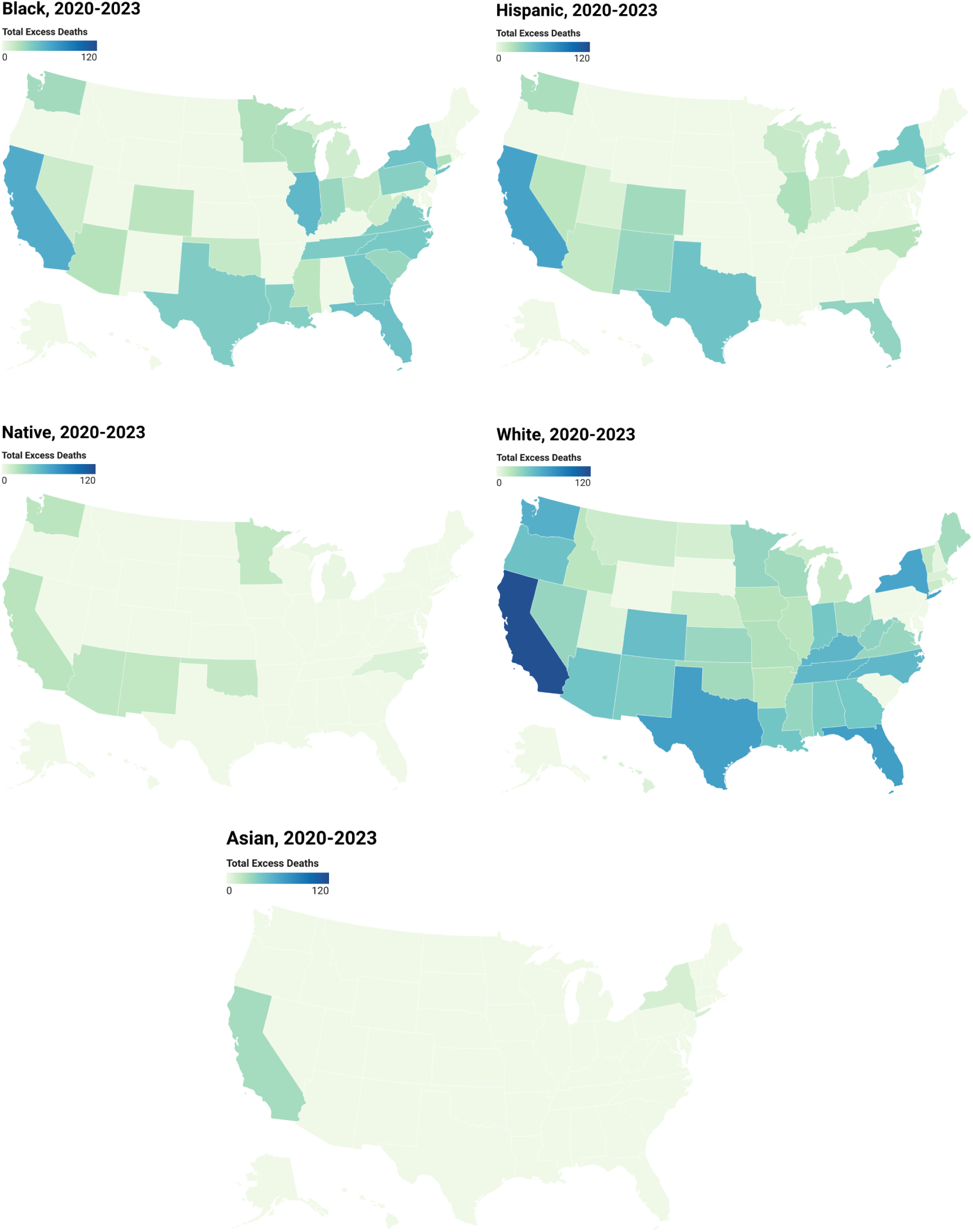
Choropleth maps of square-root-transformed total excess overdose deaths (2020–2023) by state and racial group: Black (top left), Hispanic (top right), Native American (middle left), White (middle right), and Asian (bottom). Darker blues indicate higher excess deaths; lighter greens indicate lower values. High burdens appear in California for all groups, with elevated excesses also observed in Florida, Texas, and Georgia. In the Northeast (notably New York), Black and White populations experience particularly high excess mortality, while Native American and Asian groups show lower counts overall.

**Figure S7:**
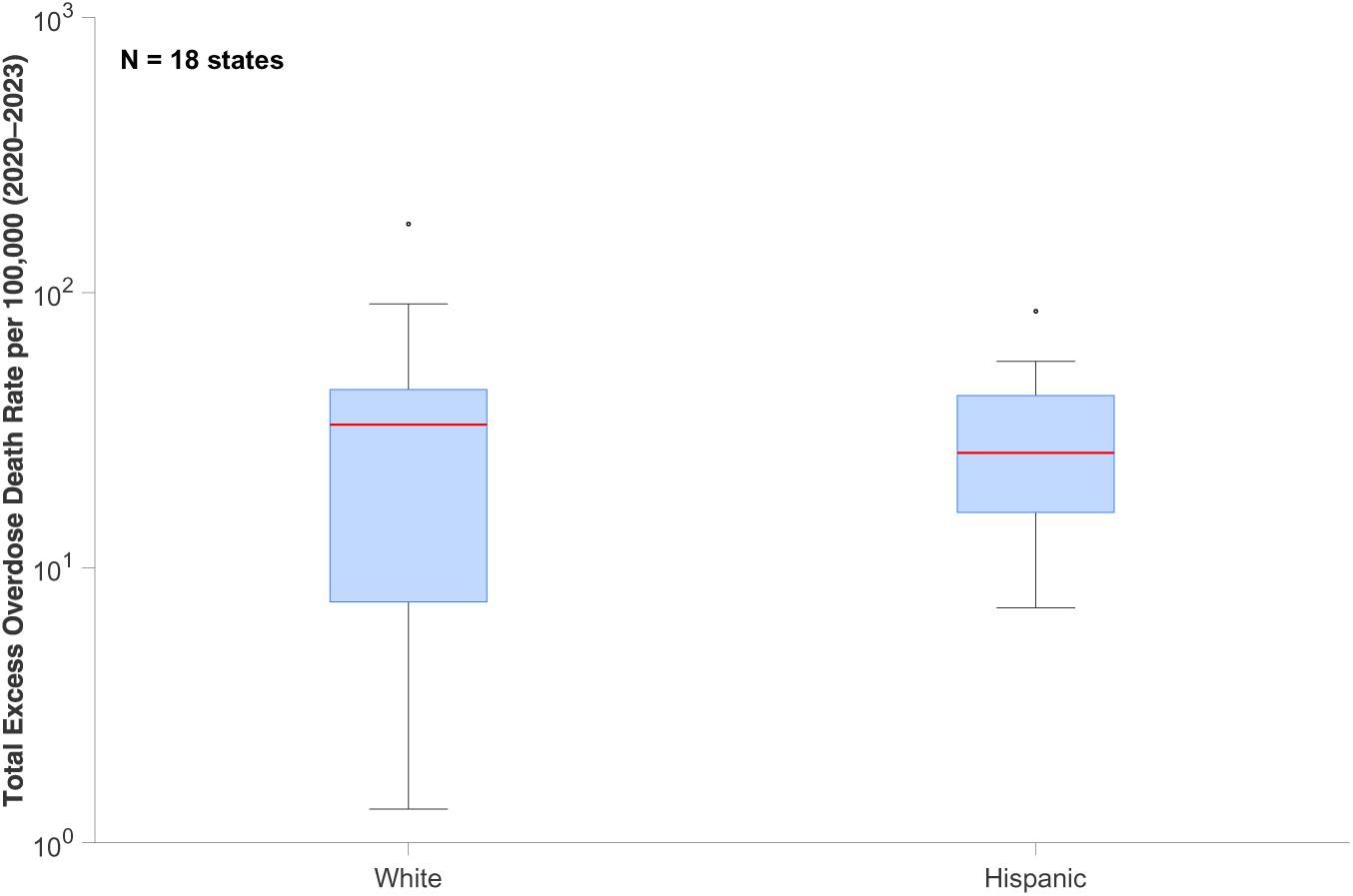
Boxplots of total excess overdose deaths rate per 100,000 (2020–2023) on a *log*_10_ scale for Hispanic and White populations. Each box aggregates state-level excess overdose death rate estimates for states with available data in both groups, i.e., (N=18 states). The Hispanic distribution shows a lower median and narrower interquartile range, whereas the White population exhibits a higher median excess and a wider spread, indicating greater variability and more extreme high-end outliers in excess deaths among the White population.

**Figure S8:**
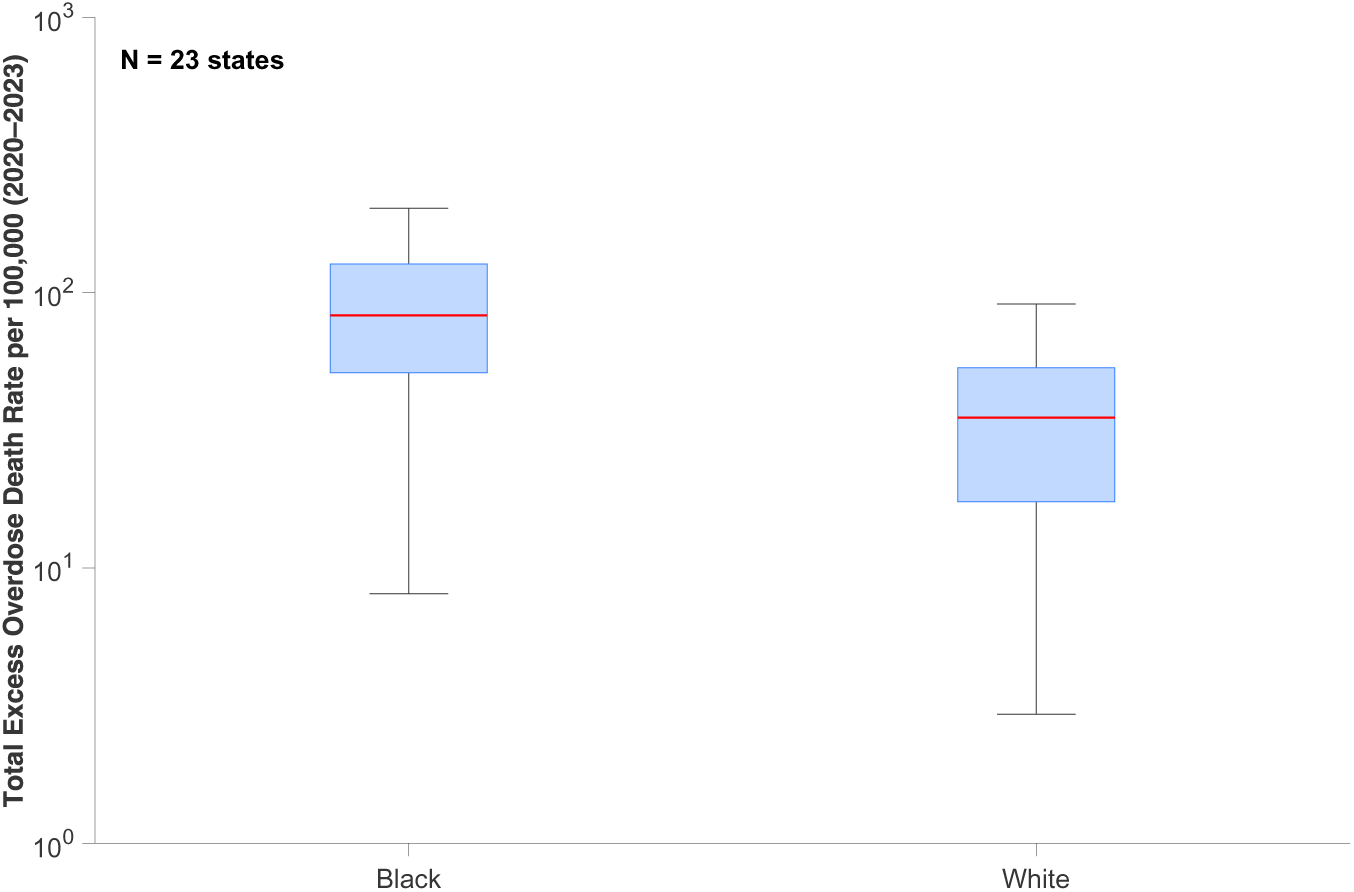
Boxplots of total excess overdose death rate per 100,000 (2020–2023) for Black and White, on a *log*_10_ scale. Each box aggregates state-level excess overdose death rate estimates for states that report data for both racial groups, i.e., (N=23 states). Red lines denote median excess deaths; whiskers extend to 1.5×IQR.

**Figure S9.**
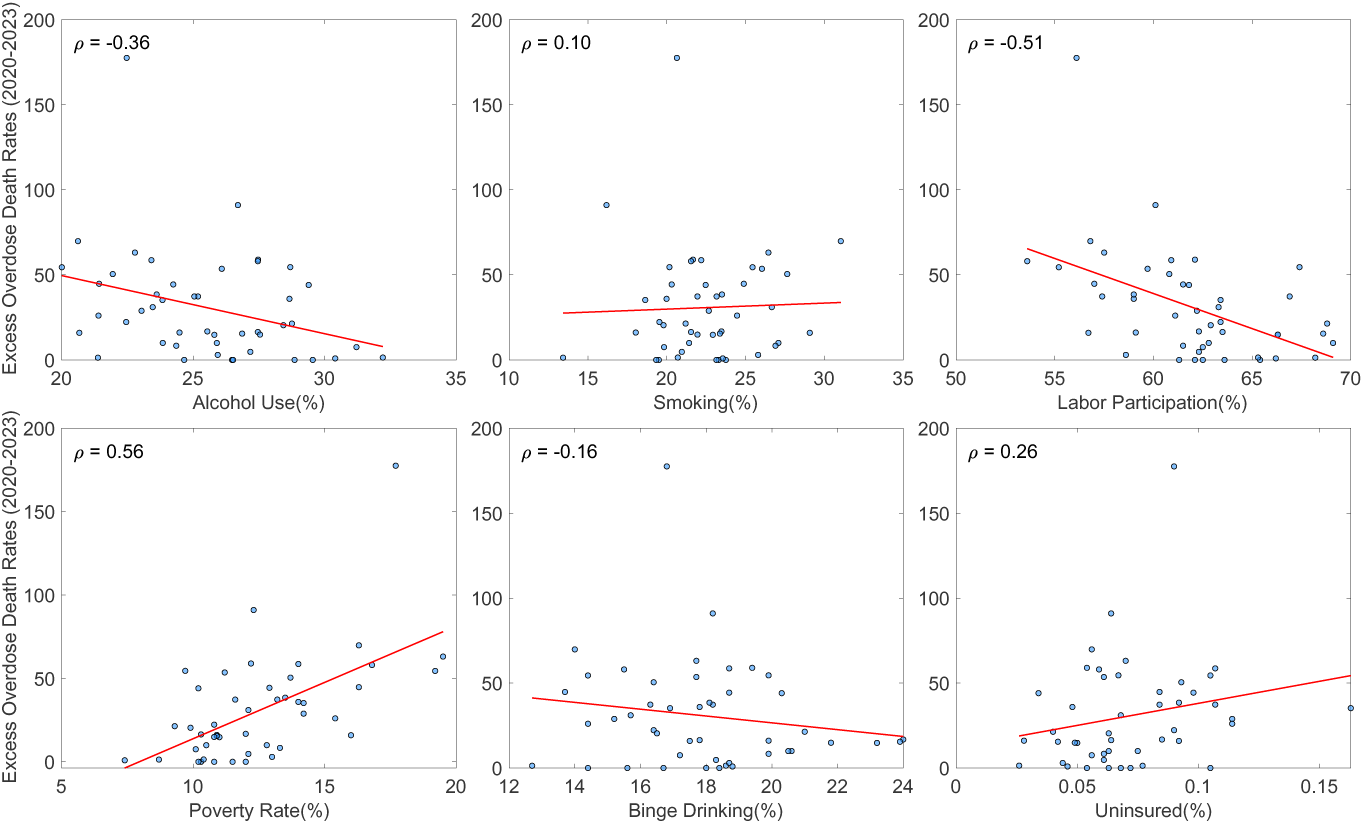
: Correlation between excess overdose mortality rates (per 100,000 population, 2020–2023) and state-level predictors among the White population. Scatterplots display bivariate associations with alcohol use, smoking, labor force participation, poverty rate, binge drinking, and uninsured rate. Each panel includes the Spearman correlation coefficient (*ρ*) and fitted regression line, illustrating the direction and strength of association between predictors and excess overdose mortality.

**Figure S10.**
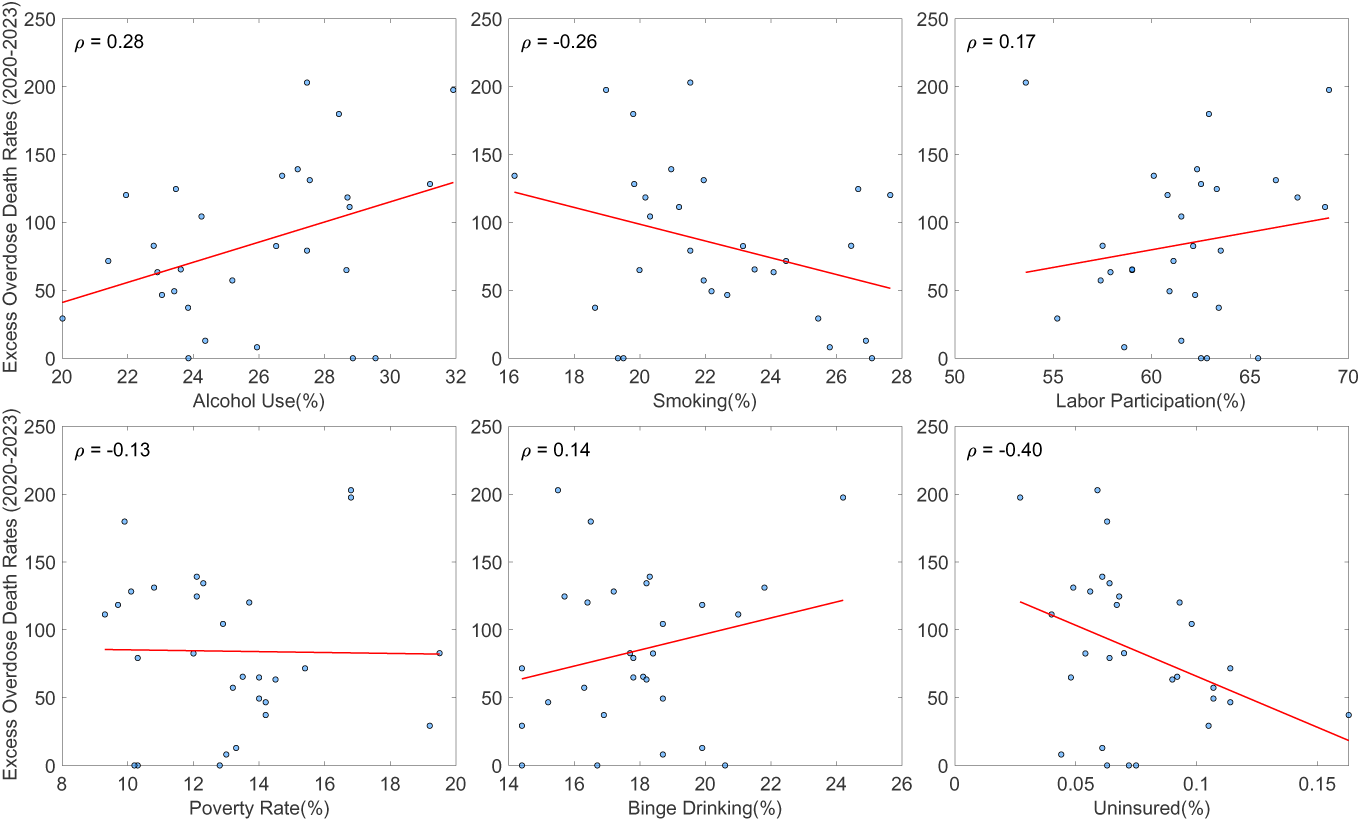
: Correlation between excess overdose mortality rates (per 100,000 population, 2020–2023) and state-level predictors among the Black population. Scatterplots display bivariate associations with alcohol use, smoking, labor force participation, poverty rate, binge drinking, and uninsured rate. Each panel includes the Spearman correlation coefficient (*ρ*) and fitted regression line, illustrating the direction and strength of association between predictors and excess overdose mortality.

**Figure S11.**
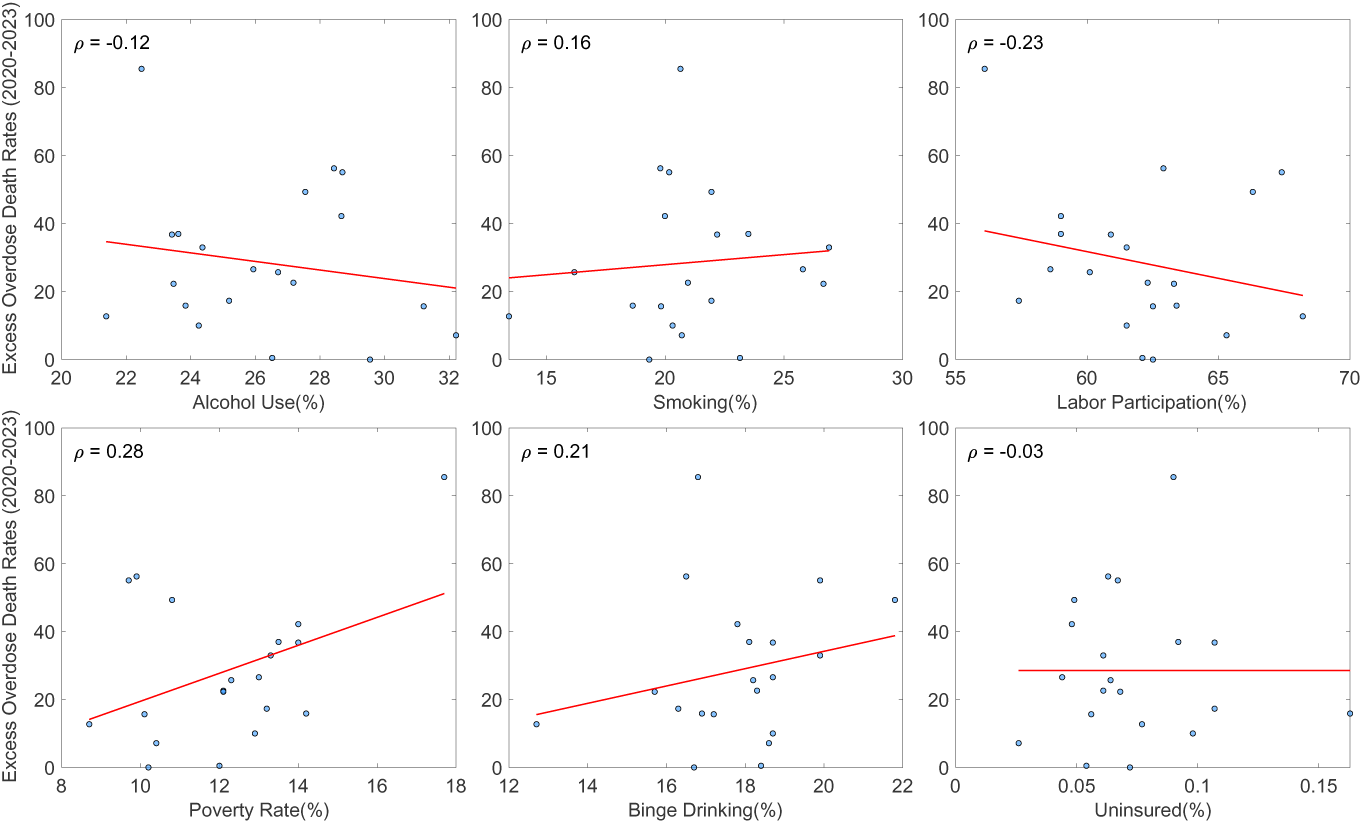
: Correlation between excess overdose mortality rates (per 100,000 population, 2020–2023) and state-level predictors among the Hispanic population. Scatterplots display bivariate associations with alcohol use, smoking, labor force participation, poverty rate, binge drinking, and uninsured rate. Each panel includes the Spearman correlation coefficient (*ρ*) and fitted regression line, illustrating the direction and strength of association between predictors and excess overdose mortality.

**Figure S12.**
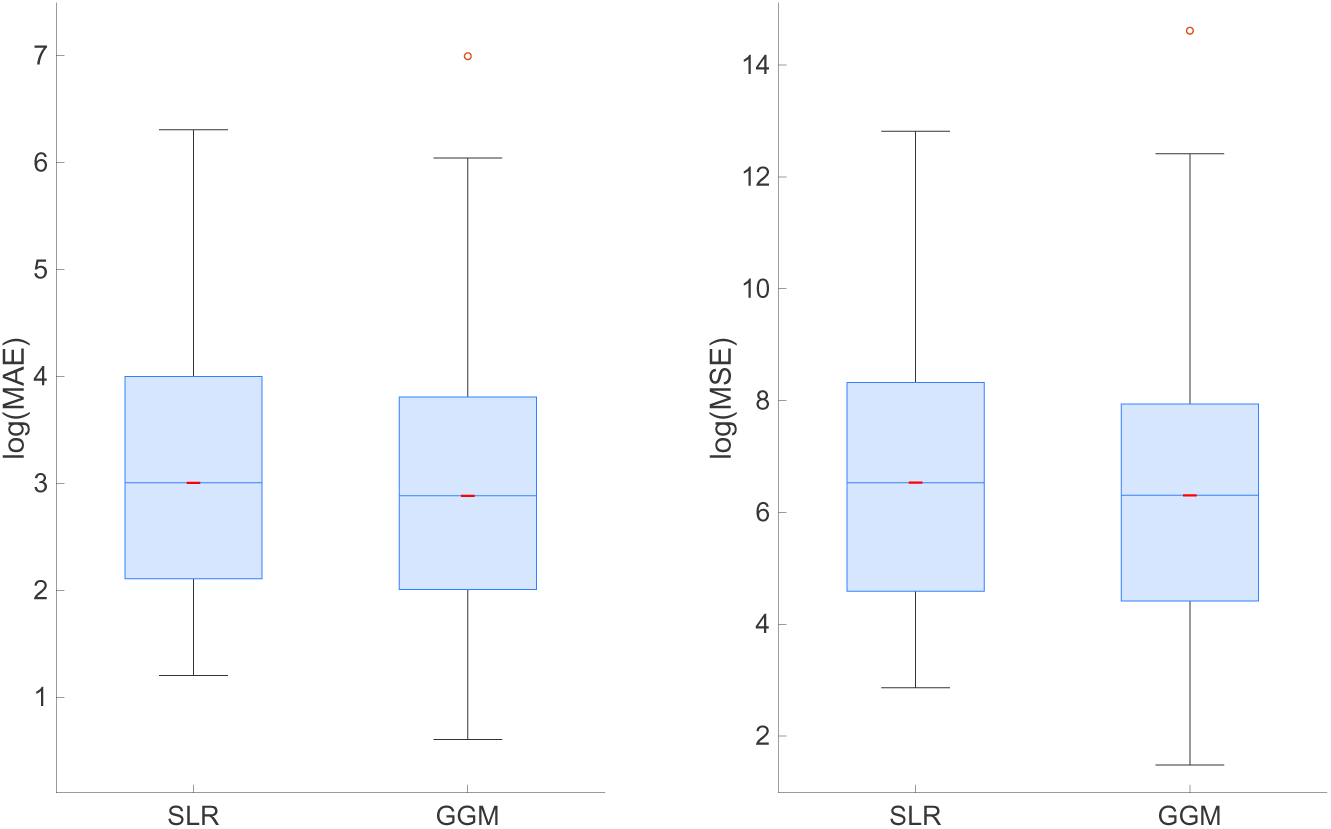
: Box plots of the log-transformed calibration performance for Simple Linear Regression (SLR) and the Generalized Growth Model (GGM) across all state–race combinations. The left panel shows the distribution of log(MAE), and the right panel shows log(MSE). Red lines indicate medians, whiskers extend to 1.5 × IQR, and outliers are plotted individually. GGM exhibits a lower median and tighter spread in both MAE and MSE compared to SLR.

